# From microbes to milestones: Gut bacterial abundances and functional pathways associate with neurodevelopment following preterm birth

**DOI:** 10.1101/2025.09.03.25334925

**Authors:** Kadi Vaher, Aisling Kenny, Paula Lusarreta Parga, Lorena Jiménez-Sánchez, Helen Turner, Rebekah Smikle, Amy Corrigan, Hilary Cruickshank, Magda Rudnicka, Sue Fletcher-Watson, Debby Bogaert, James P Boardman

## Abstract

The early life gut microbiome has been suggested to be a potential driver of neurocognitive development. Evidence for this relationship in preterm children, who are at increased risk of both gut microbiome disruptions and neurodevelopmental impairment, is scarce. In a sample of 73 very preterm infants drawn from a prospective birth cohort, we assessed associations between the neonatal gut microbiome and neurodevelopment at 9 months and 2 years. The gut microbiome was profiled from stool samples collected prior to NICU discharge using shotgun metagenomics. By taking a consensus-based analytic approach, we found strong evidence for associations between the abundances of several gut bacterial species and measures related to autistic traits (e.g. *Klebsiella spp.*), socio-emotional development, including temperament (e.g. *Enterobacter cloacae complex, Veillonella parvula*), and executive functioning (*Clostridium perfringens*). The abundances of functional modules involved in gut-brain signalling, particularly those involved in histamine, tryptophan and quinolinic acid metabolism, were associated with measures related to executive functioning and cognitive-behavioural flexibility. This study provides evidence that the neonatal gut microbiome composition may affect longer-term neurodevelopmental profiles following preterm birth, particularly those related to socio-emotional development, autistic traits and executive functioning.

## 1 Introduction

The first 1000 days of life - from conception to two years - constitute a critical period for neurodevelopment, characterised by neurogenesis, synaptogenesis, myelination, rapid brain growth, and the establishment of key neural circuits underlying cognitive, emotional, and behavioural functioning^1,2^. During this time, the developing brain demonstrates plasticity in network formation rendering it highly sensitive to both internal and external influences. Physiological perturbations can have long-lasting effects on neurodevelopmental trajectories and impact lifelong cognitive and mental health outcomes. Preterm birth (birth before 37 weeks of gestation), and very preterm birth in particular (birth <32 weeks), can interfere with the dynamic processes occurring during this crucial window of development, shaping neurodevelopment across the lifespan.

Preterm children often experience difficulties with cognitive and executive functioning, language abilities, motor skills, social interactions and emotion regulation^3–5^, and they are two to four times more likely to be diagnosed with autism^6,7^. Nevertheless, considerable heterogeneity exists within this population, and not all preterm infants experience these neurodevelopmental outcomes. This suggests that a lower gestational age (GA) alone is not sufficient to drive neurocognitive differences in children born preterm. Instead, emerging evidence highlights the impact of several, often intertwined exposures associated with preterm birth, such as infections, stress exposure and sub-optimal nutrition, which increase an infant’s likelihood for neurodevelopmental challenges^8,9^. Notably, variations in early life gut microbiome composition may contribute to the heterogeneity of neurodevelopmental outcomes observed in preterm children.

The term *gut microbiome* encompasses all microorganisms inhabiting the gastrointestinal tract, along with their collective genetic material. Here, we refer more specifically to the bacterial inhabitants of the gut microbiota (and their functional capacity), which represent the microbiome component that is highest in biomass^10^ and has been most extensively studied to date. Gut bacteria are known to play a key role in several essential biological processes such as metabolism and immune functions^11–13^, but growing evidence of their contribution to gut-brain communication and, subsequently, their influence on brain development^14–17^, may be particularly relevant in the context of preterm birth.

Associations between the early life gut microbiome and later cognitive and behavioural outcomes have been observed in typically developing children (see Vaher et al., 2022^18^ for review). For example, greater *Bacteroides* abundance within the first year has been positively correlated with language outcomes at 2–3 years of age^19–21^ and negatively with motor skills^20^, whereas early life *Bifidobacterium* abundance has shown positive associations with motor skills^22^, and infant temperament, particularly regulation/orienting and extraversion/surgency dimensions^23–26^. However, the specific microbiome features reported to associate with neurodevelopmental outcomes, including alpha (within-sample) and beta diversity (between-sample) and bacterial abundances, vary widely, which is likely due to differences in study methodology, such as the timing of microbiome and outcome assessments, and statistical analyses, as well as potential age- and sex-specific relationships^18,22,23,27,28^.

The microbiota–gut–brain axis has also been increasingly implicated in autism spectrum disorder (ASD), though findings are similarly varied. Several studies have reported compositional differences in the gut microbiota of autistic children, including decreased *Bifidobacterium* and increased *Klebsiella* and *Clostridium* species as early as five months of age, in infants having increased likelihood of later ASD diagnosis^29^. Systematic reviews and meta-analyses have documented altered abundances of *Bacteroides*, *Clostridium*, and *Bifidobacterium*^30–34^, while differences in alpha and beta diversity^29,30,35^ and disruptions in short-chain fatty acid (SCFA) metabolism^35,36^ have also been observed. However, these associations must be interpreted with caution, as it remains unclear whether microbiome alterations precede and contribute to ASD development or arise secondarily due to ASD-related behaviours, dietary preferences, or other environmental factors. Together this increasing body of literature suggests a potential role for the gut microbiome in brain development.

Nevertheless, research examining microbiome–neurodevelopmental associations specifically in preterm infants remains limited, though the importance of gut microbes in shaping brain development after preterm birth is increasingly acknowledged^37,38^. Knowing these relationships is important given that preterm infants are frequently exposed to microbiome-altering factors, such as caesarean section delivery, antibiotics, complex feeding regimens, and prolonged hospitalisation^39^, which may increase the incidence of microbiome-driven neurodevelopmental differences in this population. The gut microbiome of preterm infants follows a patterned progression, initially with high abundances of *Staphylococcus*, followed by its gradual replacement with *Enterococcaceae, Enterobacteriaceae*, and *Bifidobacteriaceae,* with a strong influence of birth GA^40–43^. Compared to their term-born peers, preterm infants are often characterised to have reduced abundance of *Bifidobacterium* and increased abundance of potentially pathogenic bacteria such as *Escherichia* and *Klebsiella*^44^. Many neonatal units routinely prescribe probiotics to prevent necrotising enterocolitis and sepsis in preterm infants^45^. However, the clinical efficacy of these probiotics remains uncertain, particularly in the most vulnerable extremely preterm infants (birth <28 weeks), safety concerns have been raised, and the evidence on the potential impact of probiotics on neurodevelopment in this population is limited^46–48^. This variation in microbiome modification in clinical practice underscores the critical importance of understanding how the neonatal gut microbiome contributes to neurodevelopmental outcomes following preterm birth.

Some previous studies have suggested that microbiome differences in the preterm population may be associated with neurostructural variation. For example, severe parenchymal brain injury was shown to associate with lower abundances of *Enterococcus*, *Escherichia/Shigella*, and *Streptococcus* in the early days of life, and higher abundances of both *Klebsiella*—a potentially pathogenic genus—and *Bifidobacterium* at 4–6 weeks of age^49^. In the larger preterm infant cohort from which this study subset is derived, we previously demonstrated that the abundances of gut bacterial species, especially *Escherichia coli* and *Klebsiella spp.*, associated with several MRI features of encephalopathy of prematurity such as deep grey matter microstructure^50^.

Associations with neurocognitive outcomes following preterm birth have also been observed in the few available studies. A seminal study utilising data from the EPIFLORE/EPIPAGE cohort found that 4-week-old very preterm infants with gut microbiome profiles characterised by high abundances of *Enterococcus* or *Staphylococcus* subsequently had higher odds of death or neurodevelopmental delay compared to infants with microbiome compositions high in *Escherichia/Shigella*^51^. Furthermore, 2-year neurodevelopmental scores from the Ages and Stages Questionnaire showed weak negative correlations with *Staphylococcus* abundance and weak positive associations with the abundances of *Escherichia*. Similarly, Abrahamsson *et al*^52^ reported that neurodevelopmental impairment-associated microbial signatures in the first month after birth mostly included higher abundances of *Escherichia/Shigella* and *Enterococcus* together with lower abundances of *Enterobacter*. In contrast, a smaller study found that reduced *Bifidobacterium* levels in the first month after birth associated with neurodevelopmental impairment at 2 years of age^53^. Sarkar *et al* report positive correlations between early gut alpha diversity and neurodevelopment in preterm infants, and distinct bacterial networks in children with typical development versus those referred for neurocognitive evaluation^54^. Two additional studies have shown dynamic changes in the way gut microbiome composition over the course of NICU stay correlates with behaviour at NICU discharge^55,56^. Collectively, these findings suggest that early gut microbiome composition in preterm infants may influence later brain and neurocognitive development, though considerable uncertainty remains regarding the specific bacterial biomarkers involved.

In this work, we leveraged a richly phenotyped cohort of very preterm infants^57^ with the aim to uncover the associations between the neonatal gut microbiome diversity and community composition and neurobehavioural outcomes in the first two years of life^58^.

## 2 Materials and Methods

Given the substantial variability and mixed findings in previous literature, as well as our focus on very preterm infants who have been underrepresented in this type of research, we adopted an exploratory approach to comprehensively assess potential relationships across several neurodevelopmental domains. Given the high analytic freedom, we adopted a consensus-based, multiverse analytical approach, which was preregistered prior to analyses^58^. We have included descriptions of deviations from this analysis plan below. We followed the Strengthening The Organization and Reporting of Microbiome Studies (STORMS) checklist^58^ in describing the methodology and reporting results (Supplementary table 1).

### 2.1 Participants

Participants were very preterm infants born at <33 completed weeks’ gestation recruited to the Theirworld Edinburgh Birth Cohort (TEBC), a longitudinal study designed to investigate the effects of preterm birth on brain structure and long-term developmental outcomes^57^. Participants were recruited and faecal samples for gut microbiome profiling were sampled at the Royal Infirmary of Edinburgh, UK, between 2016-2021. Exclusion criteria were death during neonatal period, major congenital malformations, chromosomal abnormalities and congenital infection. The study was conducted according to the principles of the Declaration of Helsinki, and ethical approval was obtained from the UK National Research Ethics Service (South East Scotland Research Ethic Committee 16/SS/0154). Informed consent was obtained from a person with parental responsibility for each participant.

All infants were cared for in the Neonatal unit of the Simpson Centre for Reproductive Health, Royal Infirmary of Edinburgh, following standardised feeding and antibacterial guidelines (see previous work^50,59^ and Supplementary materials for details). Preterm infants admitted to the NICU in the Simpson Centre for Reproductive Health are not routinely administered any pro- or prebiotic supplements. Clinical data was collected from antenatal and neonatal electronic patient records (for definitions see Supplementary table 2).

The participants of this work were preterm infants who had their gut microbiome samples obtained prior to discharge from the NICU and who subsequently had any neurodevelopmental outcome data available at 9 months and/or 2 years; see below and in Supplementary materials for details.

### 2.2 Gut microbiome

Faecal sample collection, bacterial DNA extraction and shotgun metagenomic sequencing are described in detail in our previous publication^50^ and in Supplementary materials. In brief, bacterial DNA from stool samples was extracted using a protocol that involves phenol/bead beating in combination with the Mag Mini DNA Isolation Kit (LGC genomics, Germany), previously known as ‘*Agowa* Mag Mini DNA Isolation Kit’^60^; shotgun metagenomic sequencing was performed on the NovaSeq 6000 platform (Illumina); taxonomic and functional profiling were carried out using MetaPhlAn (v3.1) and HUMAnN (v3), respectively^61^. Functional gene family data were grouped to KEGG orthogroups (KOs) from which gut-brain modules (GBM) were calculated^62^.

Shotgun sequencing data was available for a total of 97 preterm infants at pre-discharge^50^; two samples were excluded from the analyses due to collection at a very early postmenstrual age (<33 weeks), prior to infant transfer to another neonatal unit (see Supplementary materials for further info on sample inclusion). In total, 73 preterm infants with microbiome data had at least one neurodevelopmental outcome measure available (see below). After removing species and KOs absent in these 73 infants and those present in <5% of the sample (i.e. species only present in up to 3 samples, a total of 106 species and 3281 KOs), the dataset consists of 71 species and 4384 KOs. From the KOs we calculated a total of 41 GBMs.

### 2.3 Behavioural outcomes

We collected information about infants’ neurodevelopment at 4.5 months, 9 months and 2 years; behavioural assessments were conducted at corrected age for preterm infants.

#### Infant Behaviour Questionnaire (IBQ) at 4.5 months and 9 months and Early Childhood Behavioural Questionnaire (ECBQ) at 2 years

These parent reported questionnaires are designed to assess temperament between the ages of 3 and 12 months^63^, or 18 and 36 months^64^, respectively. From the questionnaire data, we calculated three broad dimensions of temperament: negative affectivity, surgency/extraversion, and regulation (equivalent to effortful control domain in ECBQ). Higher scores indicate higher prevalence of behaviours indicative of these traits. Note that data collected at 4.5 months was not included in the statistical analyses but was used to impute data for 10 infants for the 9 month timepoint; please see Supplementary materials for details.

#### Behaviour Rating Inventory of Executive Function, Preschool Version (BRIEF-P) at 2 years

BRIEF-P is designed to assess the range of behavioural manifestations of infant’s executive function based on parental report^65^. We calculated the Global Executive Functioning as the composite score for executive functioning, with higher scores indicative of greater executive functioning difficulties. Infants with high negativity or inconsistency ratings (n=1) were not included in the analyses.

#### Quantitative Checklist for Autism in Toddlers (Q-CHAT) at 2 years

Q-CHAT is designed to assess frequency of behaviours also observed in autism spectrum conditions in toddlers aged 18 to 30 months based on parental report and identify those who should be referred for diagnostic assessment^66^. We obtained a total Q-CHAT score, with higher scores indicative of more autistic traits.

#### Bayley Scales of Infant and Toddler Development, Third Edition (Bayley-III) at 2 years

Bayley-III^67^ was conducted as part of the routine National Health Service developmental follow-up appointment at 2 years corrected age; data was extracted from medical records for the purposes of this study. We obtained the standardised composite scores for the five domains measured with Bayley-III: cognitive, motor, language, adaptive, and socio-emotional development, with higher scores indicative of better functioning in these domains. In Bayley-III, the cognitive, motor and language domain scores are determined through clinical assessment, while adaptive and socio-emotional domains are parent reported.

### 2.4 Quantification and statistical analysis

Analyses were conducted separately for each outcome measure, given not all 73 infants had data available for every measure. This approach resulted in 13 analysis subsets (one per outcome) to maximise sample size for each measure.

For a scheme depicting the quantification and analysis flow, please see Supplementary Figure 1. We used a consensus-based/multiverse approach, using different indices of alpha and beta diversity, and several methods of differential abundance testing. All statistical analyses were performed in R v4.4.2.

#### 2.4.1 Beta diversity and PERMANOVA

For both taxonomic and functional data, we calculated both Bray-Curtis dissimilarity (using total-sum-scaled [i.e. relative abundances] species/KO tables) and Aitchison’s distance (using centred log-ratio [CLR] transformed species/KO data) using the *vegan* package^68^. Then, we used permutational analysis of variance (PERMANOVA), modelled by *adonis2* with 1000 permutations, to identify associations between overall microbiome community composition (i.e. beta diversity) and the outcomes of interest. Raw p-values were adjusted for multiple comparisons using the Benjamini-Hochberg method separately for each matrix; we considered raw p-values < 0.05 as nominally significant, and adjusted p-values (q) < 0.05 as statistically significant. In addition to the registered analysis for beta diversity, to assess the agreement of the results when using the two distance/dissimilarity measures, we calculated Pearson’s correlation coefficients and mean [range] absolute differences in the R^2^ values.

#### 2.4.2 Dimensionality reduction

As in our previous work^50^, principal coordinates analysis (PCoA) was performed on the species-level Bray-Curtis dissimilarity matrix using the function *pcoa* (*ape* package^69^). Cailliez transformation was applied to correct for negative eigenvalues^70^. We extracted 5 principal coordinates (PCos) each explaining >5% of variance in the data, cumulatively explaining approximately 50% of variance in microbiome composition data. For taxonomic and functional interpretation, Spearman correlation coefficients were calculated between the PCo scores and the relative abundances of the species and GBMs (Supplementary Figures 2-3). The community PCos derived here are similar to those derived from 16S-based sequencing^50^.

#### 2.4.3 Alpha diversity

For both taxonomic and functional data, Shannon index and number of observed species/KOs were calculated, before 5% prevalence filtering. As MetaPhlAn outputs species data in relative abundances, estimates of species read counts were first computed by multiplying the relative abundance of a species in a sample by the number of total sequences generated by that sample (following quality checks). For functional data, alpha diversity indices were calculated for the KOs in RPK units rounded to the nearest integer.

#### 2.4.4 Partial Spearman correlation

Initially, as outlined in the preregistration, we performed linear regression models to investigate associations for microbiome alpha diversity indices and community composition PCos with outcome measures. However, inspection of model diagnostic plots revealed that there were violations of the assumptions, especially linearity and normal distribution of residuals. We assessed the impact of log transformations on microbiome alpha diversity indices and PCos to improve compliance with model assumptions, however, diagnostic plots continued to indicate violations of certain assumptions Therefore, we performed (partial) Spearman correlation analysis as a nonparametric alternative to associate microbiome alpha diversity indices and PCos with outcomes. As with PERMANOVA, raw p-values were adjusted for multiple comparisons using the Benjamini-Hochberg method separately for each alpha diversity index and PCo; we considered raw p-values < 0.05 as nominally significant, and adjusted p-values (q) < 0.05 as statistically significant.

#### 2.4.5 Differential abundance testing

We used three complementary differential abundance methods to investigate associations between individual microbiome features (species and GBMs) and outcomes: MaAsLin2^71^, LinDA^72^, and a multiple linear regression with CLR-transformed abundance data (as recommended in the *Bugs as features* tutorial^73^). These methods were chosen as they provide flexible options for covariate adjustment, are widely used in the field, and are heterogenous in their assumptions and data transformation/normalisation approaches. To reduce the number of comparisons, we analysed microbial features which were present in at least 20% of the full sample of 73 infants, resulting in a set of 31 species and 28 GBMs to be analysed using differential abundance testing methods.

For MaAsLin2, we used the relative abundance feature table as the input. No further normalisation or filtering was performed, and default options were followed. Outcome measures along with relevant covariates (see next section) were specified as fixed effects.

For the linear regression modelling, 5% prevalence-filtered feature relative abundance data was CLR-transformed. Then, linear regression models were performed for each feature present in >20% of the sample and outcome measure, adjusting for relevant covariates.

For LinDA, estimates of species read counts were computed by multiplying the relative abundance of a species in a sample by the number of total sequences generated by that sample. Next, the species count data was used as input for the model; the outcome measures along with relevant covariates were specified in the formula. For functional data, RPK-based GBM data was utilised as input. LinDA was chosen to replace the originally preregistered ANCOM-BC2^74^ method. This was due to the relatively small sample size, which resulted in an ANCOM-BC2 output with many NA values, especially as model complexity increased with the inclusion of covariates.

In all differential abundance methods, Benjamini-Hochberg correction for multiple comparisons was used across the microbiome features separately for each outcome measure. Results with p < 0.05 were considered nominally significant and those with adjusted p-value (q) < 0.25 (as per the default in MaAsLin2) were considered statistically significant. To reflect the consensus-based analysis, results from the three differential abundance testing methods were collated into *evidence levels* based on concordance across methods. Namely, we defined *strong evidence* if there were statistically significant associations (q < 0.25) between a bacterial feature and an outcome measure in at least two of the three methods used, and *moderate evidence* if there was a statistically significant association between a bacterial feature and an outcome measure in at least one of the three methods used along with a nominally significant (p < 0.05) association in at least one or more of the three methods used. *Weak evidence* was defined as nominally significant (p < 0.05) associations in at least two of the three methods used. Any other associations that only arose in one method are not discussed.

To further investigate consensus and alignment between the three different methods, the raw effect sizes were converted to z-scores and Pearson correlation coefficients were computed between the z-scored effect sizes for each pair of the three differential abundance testing methods.

#### 2.4.6 Partial least squares regression

To further evaluate the robustness of the microbiome-outcome relationships, in addition to the above-described preregistered analyses, we performed sparse partial least squares regression (sPLS) analysis using the *mixOmics* package^75^. These analyses were performed separately for all behavioural outcome measures for which at least one microbiome feature was found to be associated at the strong or moderate evidence level in differential abundance analyses. The predictor was the CLR-transformed and 20%-prevalence filtered feature abundance table (i.e. the same 31 species and 28 GBMs that were used in differential abundance analyses). Both the feature table and behavioural variable were subset to those participants for whom this specific behavioural data was available. We used mean absolute error and cross-validation implemented in the *tune.spls* function to identify the optimal number of components and microbiome features. We report the proportion of variance explained by the resulting components, loadings of the microbiome features to the components, and the correlation between the selected components and the behavioural measure. Finally, we extracted the resulting microbiome components and performed linear regression analyses with the behavioural measure as the outcome and selected microbiome feature component(s) along with all covariates (see next) as the predictors to ensure the correlation was not due to confounding variables impacting either microbiome or behavioural outcomes.

#### 2.4.7 Covariate identification and adjustment

In univariable analyses, associations for a range of potential confounders with microbiome alpha diversity indices, community PCos, and outcome measures were evaluated (see section Covariate identification in Supplementary materials for more information). Depending on the distribution of microbiome features and outcome scores, for categorical covariates, we performed two-sample t-tests or Mann-Whitney U-tests, and for continuous variables, we performed Pearson or Spearman correlations. In all analyses described above, we performed multivariable models where we adjusted for any covariates that were suggestively (unadjusted p<0.1) associated with at least one microbiome feature (community PCo or alpha diversity index) and at least one neurodevelopmental outcome measure. The final list of covariates included in all fully adjusted models were: GA at birth, postmenstrual age (PMA) at sample, antibiotics < 72h after birth, birthweight z-score, high vs low proportion of exclusive breast milk days during NICU stay, infant sex, maternal BMI at pregnancy booking, maternal age and the Scottish Index of Multiple Deprivation (SIMD) quintile. For description and coding of the covariates, please see Supplementary Table 2. Maternal education and postnatal depression score also met our definition of confounder, but these variables were omitted because they were missing for >5% of participants.

Given that both antibiotic exposure and breastfeeding are strong drivers of infant gut microbiome composition in early life^39^, we performed another, partially adjusted model, excluding the breast milk and antibiotic covariates. This was to assess whether including these strong microbiome determinants in our main fully adjusted model could constitute over-adjustment and potentially mask relevant biological associations.

In the main text, we report results that have been adjusted for all covariates. Results arising from univariable (i.e. unadjusted), age-adjusted (i.e. adjusted for GA at birth and PMA at sample only) and partially adjusted models (i.e. without antibiotic and breast milk variables) are presented in Supplementary materials.

#### 2.4.8 Sensitivity analyses

In case of any statistically significant associations between the gut microbiome features and 9-month temperament dimensions measured with IBQ, sensitivity analyses were performed excluding the 10 infants with imputed scores (see Supplementary materials) and adjusting for all covariates.

#### 2.4.9 Data availability

All data generated in this study are stored at the University of Edinburgh. The data are not part of an open repository due to the terms of the informed parent/guardian consent, which stipulates that the use of anonymised data is for studies of perinatal health that have been approved by regulatory bodies. All raw shotgun data and their derivatives used for analysis alongside participant clinical data, sample metadata, and outcome data used in this work are deposited in Edinburgh DataVault^76^ (https://doi.org/10.7488/e65499db-2263-4d3c-9335-55ae6d49af2b). Requests for access will be considered under the study’s Data Access and Collaboration policy and governance process (https://www.ed.ac.uk/centre-reproductive-health/tebc/about-tebc/for-researchers/data-access-collaboration). Code used for the data analysis in this paper is available on GitLab (https://git.ecdf.ed.ac.uk/jbrl/microbiome-and-developmental-outcomes). Specific packages and software used are described in Supplementary table 3. Additional information is available from the corresponding author upon request.

## 3 Results

### 3.1 Participant characteristics and neurodevelopmental outcomes

73 preterm participants had available data for shotgun metagenomic sequencing prior to NICU discharge (roughly term-equivalent age [average 36.57 postmenstrual weeks]), and at least one neurodevelopmental outcome measure (Table 1). None of the participants had necrotising enterocolitis. The sample sizes, distributions and intercorrelations of the outcome measures are presented in Figure 1. As expected, all Bayley-III domains showed positive correlations between one another; Q-CHAT scores negatively correlated with all Bayley-III domains; BRIEF-P had a negative correlation with Bayley-III cognitive domain and with the 2-year effortful control temperament dimension; Bayley-III adaptive behaviour scale also positively correlated with regulation/effortful control temperament dimension at 9 months and 2 years. The temperament measures at the two timepoints appeared less correlated, potentially reflecting a developmental change; yet there were moderate positive correlations in surgency and effortful control between 9 months and 2 years.

**Figure 1.**
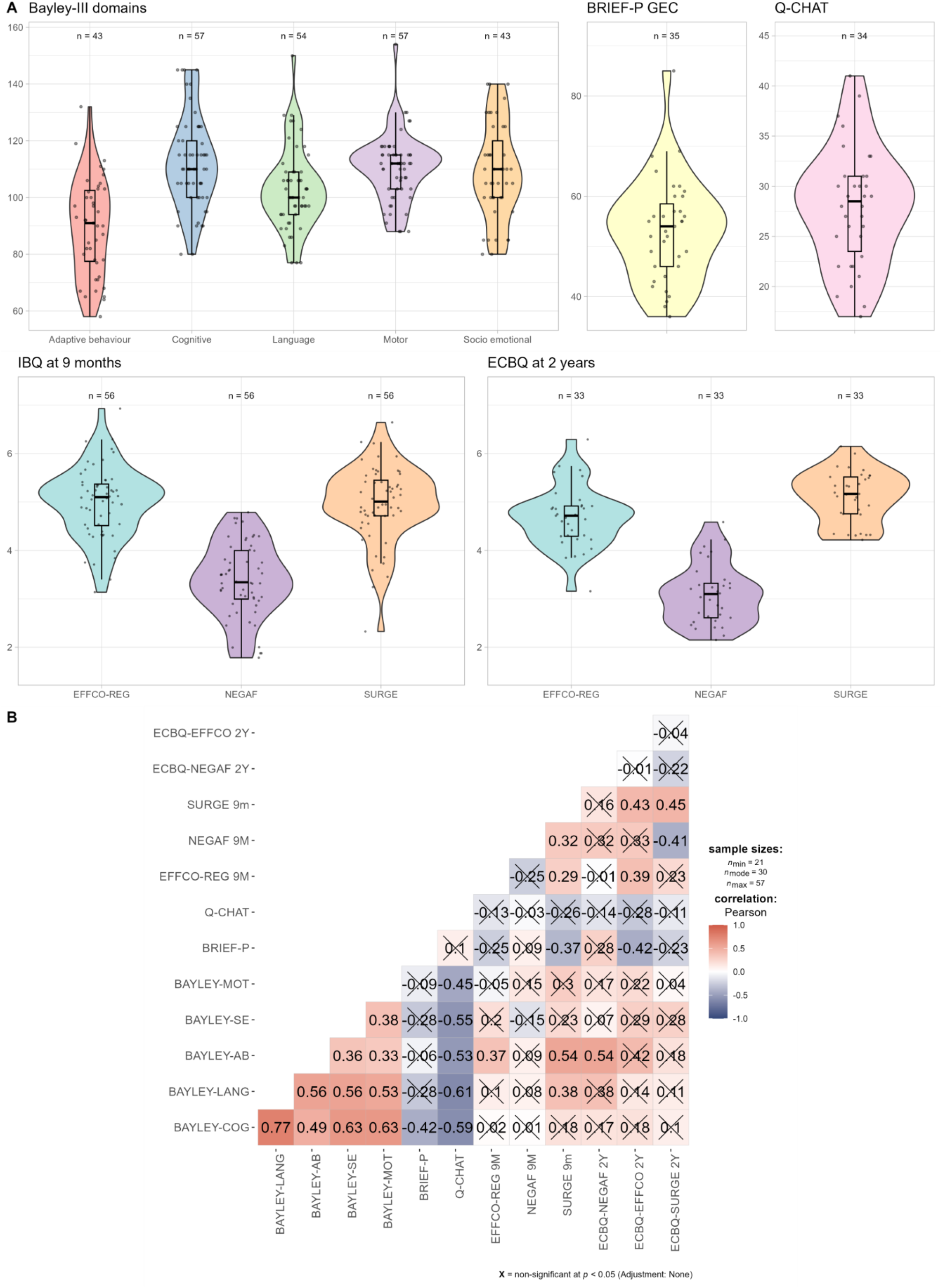
Distributions (A) and Pearson correlations (B) of neurodevelopmental outcome measures. Crossed out boxes indicate statistically non-significant correlations. BRIEF-P = Behavior Rating Inventory of Executive Function, Preschool, GEC = global executive composite, Q-CHAT = Quantitative Checklist for Autism in Toddlers; IBQ = Infant Behavior Questionnaire; ECBQ = Early Childhood Behavior Questionnaire; EFFCO-REG = effortful control/regulation, NEGAF = negative affectivity, SURGE = surgency, AB = adaptive behaviour, LANG = language, COG = cognitive, MOT = motor, SE = socio-emotional.

**Table 1.** Sample characteristics.

| Characteristic | N = 73* |
| --- | --- |
| **Infant characteristics** |  |
| GA at birth, weeks | 29.71 [22.14 to 32.86] |
| Postnatal age at sample, days | 49 [15 to 119] |
| PMA at sample, weeks | 36.57 [33.57 to 43.86] |
| PMA at discharge, weeks | 37.57 [34.43 to 45.86] |
| Male infants | 41 (56%) |
| Birthweight, grams | 1,320 (434) |
| Birthweight z-score | 0.246 [-2.520 to 2.141] |
| Vaginal delivery | 22 (30%) |
| NICU inpatient days | 63 (31) |
| Antibiotics <72h after birth | 59 (81%) |
| Antibiotics >72h after birth | 38 (52%) |
| Breastmilk exposure ≥ 75% inpatient days | 38 (52%) |
| Proportion of days receiving exclusive breastmilk in NICU | 0.778 [0.000 to 1.000] |
| Bronchopulmonary dysplasia | 21 (29%) |
| Sepsis (early- and/or late-onset) | 17 (23%) |
| **Maternal characteristics** |  |
| Age, years | 33 (5) |
| Body mass index | 25.5 [16.6 to 46.6] |
| SIMD quintile |  |
| 1 | 12 (16%) |
| 2 | 9 (12%) |
| 3 | 14 (19%) |
| 4 | 10 (14%) |
| 5 | 28 (38%) |

\*Data presented as mean (SD) for continuous, normally distributed variables; median [range] for continuous, not normally distributed variables; n (%) for categorical variables. PMA = postmenstrual age, GA = gestational age, NICU = neonatal intensive care unit. SIMD = Scottish Index for Multiple Deprivation (1 = most deprived, 5 = least deprived).

### 3.2 Microbiome correlations with behavioural outcomes

#### 3.2.1 Community level associations

Microbiome community compositions of the infants included in this study are visualised in Figure 2A. Prior to NICU discharge, the gut microbiome of these infants were dominated by high levels of *Bifidobacterium* spp. (*B. breve, B. longum*), *E. coli*, *E. faecalis* or *Klebsiella* spp. *(K. oxytoca, K. michiganensis, K. pneumoniae*).

**Figure 2.**
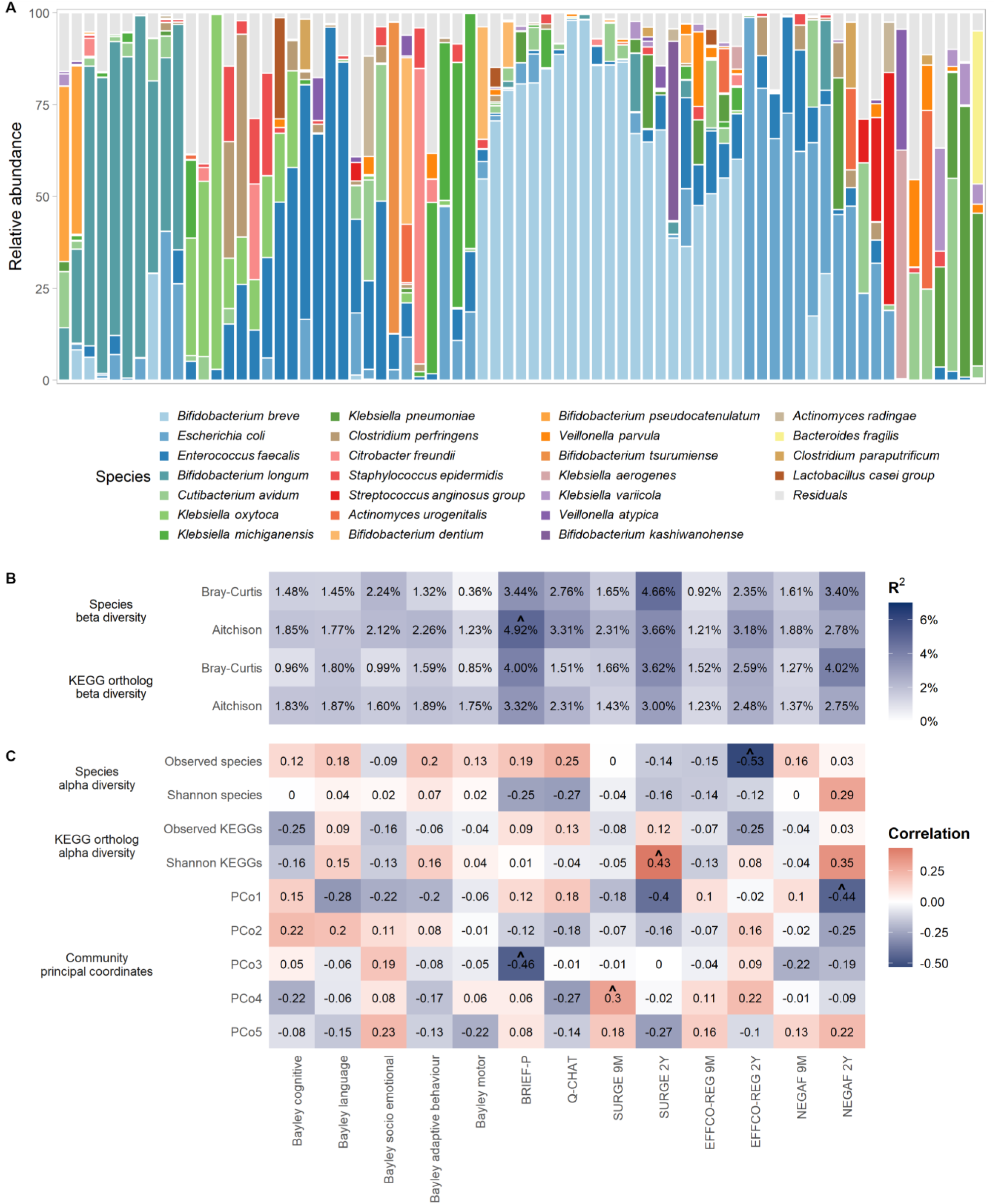
Outcomes in preterm infants were minimally associated with overall microbiome diversity and compositional variance. (A) Relative abundances of the 25 most abundant species identified across the dataset are visualised per sample, with all other species grouped together as residuals. Samples are ordered based on hierarchical clustering of the Bray-Curtis dissimilarity matrix using complete linkage. (B) Heatmap of permutational analysis of variance (PERMANOVA) results showing associations between outcome measures and the gut bacterial beta diversity calculated on species and KEGG ortholog data. Significance of PERMANOVA was based on 1,000 permutations. (C) Heatmap of partial Spearman correlation coefficients showing associations between gut microbiome alpha diversity calculated from species and KEGG ortholog data, and gut microbiome community principal coordinates (PCo) derived from principal coordinates analysis applied on species-level Bray-Curtis dissimilarity matrix. All models were adjusted for GA at birth, PMA at sample, antibiotic exposure < 72h of life, birthweight z-score, high vs low proportion of exclusive breast milk days during NICU stay, infant sex, Scottish Index of Multiple Deprivation quintile, maternal BMI at pregnancy booking, and maternal age. P-values were adjusted for multiple comparisons using the Benjamini-Hochberg (BH) method; asterisks denote statistical significance (^p < 0.05 [nominally significant], *q < 0.05 [statistically significant after adjustment for multiple tests]).

We found minimal evidence that the overall gut microbiome structure, including taxonomic and functional alpha or beta diversity, or community composition PCos, correlate with behavioural outcomes (Figure 2B-C; Supplementary tables 4-7), regardless of covariate adjustment strategy. Some nominally significant associations were observed but these did not survive adjustment for multiple comparisons. For example, BRIEF-P explained close to 5% of the variance in microbiome taxonomic community composition (when measured with Aitchison’s distance) and had a moderate negative correlation with PCo3 (partial Spearman ρ=-0.46), but these associations were no longer statistically significant following FDR correction. For beta diversity analysis, we observed good concordance/agreement in results obtained using the two different distance/dissimilarity measures: the Pearson correlation of the R^2^ values obtained using Aitchison vs Bray-Curtis was 0.832 for species and 0.867 for KOs, and the mean [range] absolute differences between the R^2^ values were 0.64% [0.12% – 1.48%] for species and 0.53% [0.07% – 1.27%] for KOs.

#### 3.2.2 Species and functional modules: Associations with autistic traits, socio-emotional development and executive functioning

The results of the three differential abundance testing methods were overall well aligned with one another. For species-level analyses, we observed Pearson correlations of r = 0.86, r = 0.90 and r = 0.94 between the effect sizes from LinDA and MaAsLin2, LinDA and linear regression, and MaAsLin2 and linear regression, respectively. For GBM-level analyses, the respective Pearson correlation coefficients were r = 0.92, r = 0.89, r = 0.88.

Species-level differential abundance testing showed *strong evidence* for a total of six species-outcome associations in the fully adjusted model. In particular, the abundances of *Klebsiella* spp. (*K. pneumoniae, K. quasipneumoniae, K. variicola)* were positively, and *Cutibacterium avidum* and *Staphylococcus haemolyticus* negatively associated with Q-CHAT scores (Figure 3; see Supplementary tables 8-11 for coefficients, standard errors and p-values of the three methods for unadjusted, age-adjusted, fully adjusted and partially adjusted models). The abundance of *Enterobacter cloacae complex* was positively associated with the Bayley-III socio-emotional development domain.

**Figure 3.**
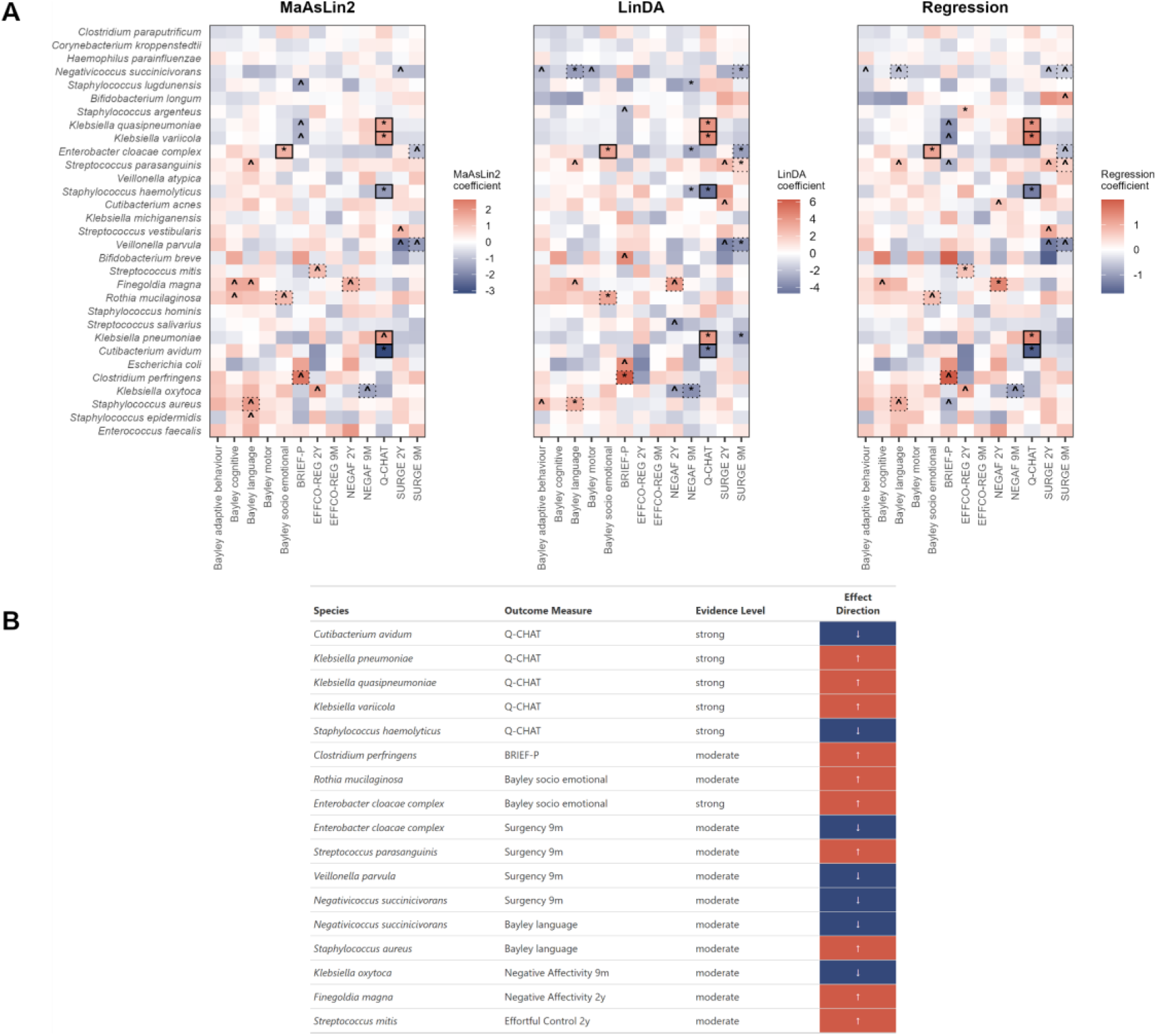
Gut microbiome species relative abundances associate with socio-emotional development. (A) Heatmaps of the results of differential abundance analyses showing associations between species abundance and outcome measures of interest. Symbols denote significance level: *statistically significant (q < 0.05 after adjustment for multiple comparisons), ^nominally significant (p < 0.05 prior to adjustment). Boxed cells denote evidence level: solid boxes indicate strong evidence level while dashed boxes indicate moderate evidence level. Species are ordered based on prevalence, with the most prevalent (i.e. missing in the smallest number of infants in the whole sample) at the bottom. (B) Table highlighting strong and moderate associations observed in the differential abundance analyses. Direction of effect is indicated by colour scheme (blue for negative, red for positive).

*Moderate evidence* was observed for another 11 associations between relative abundances of bacterial species and outcome measures. The abundance of *Streptococcus parasanguinis* was positively and the abundances of *Negativicoccus succinicivorans*, *Veillonella parvula* and *Enterobacter cloacae complex* were negatively associated with the temperament dimension of surgency at 9 months of age. *Enterobacter cloacae complex* also strongly associated with Bayley-III socio-emotional domain as described above. Negative associations for the abundances of *Negativicoccus succinicivorans* and *Veillonella parvula* as well as positive associations for *Streptococcus parasanguinis* were also observed with surgency at 2 years of age, albeit at the *weak evidence* level (i.e. nominally significant in at least two methods but did not survive adjustment for multiple comparisons). *Negativicoccus succinicivorans* again showed moderate evidence for an association with the Bayley language domain and weak evidence for an association with Bayley adaptive behaviour.

Differential abundance testing of the GBMs revealed *strong evidence* for four GBM-outcome associations. We found positive associations for the abundances of histamine and quinolinic acid synthesis with BRIEF-P (Figure 4A-B; see Supplementary tables 12-15 for coefficients, standard errors and p-values of the three methods for unadjusted, age-adjusted, fully adjusted and partially adjusted models). Quinolinic acid synthesis and 17-β-estradiol degradation showed negative associations with the effortful control/regulation trait at 9 months and 2 years, respectively. Five associations showed *moderate evidence* for GBM abundance and outcome correlations. Notably, quinolinic acid *degradation* negatively (and quinolinic acid *synthesis* positively) associated with the BRIEF-P measure. *Clostridium perfringens* was the strongest contributor to the histamine synthesis module and it was among the top six contributors to both quinolinic acid degradation and synthesis modules (Figure 4C), all of which showed significant associations with BRIEF-P. BRIEF-P was additionally negatively associated with the menaquinone (Vitamin K2) synthesis module, and Bayley-III adaptive behaviour negatively associated with 17-β-estradiol degradation module. With regards to the GBM-outcome associations identified at the *weak evidence* level (Supplementary table 14), it is noteworthy that the *Klebsiella spp.* associated with Q-CHAT, are among the top contributors to several GBMs associated with Q-CHAT (Supplementary figure 7): acetate degradation, GABA synthesis, propionate synthesis, and the isovaleric acid synthesis (KADH pathway); whereas *Staphylococcus aureus* is the top contributor to nitric oxide synthesis GBM, both of which were positively correlated with Bayley language domain at the *moderate* and *weak* evidence levels, respectively.

**Figure 4.**
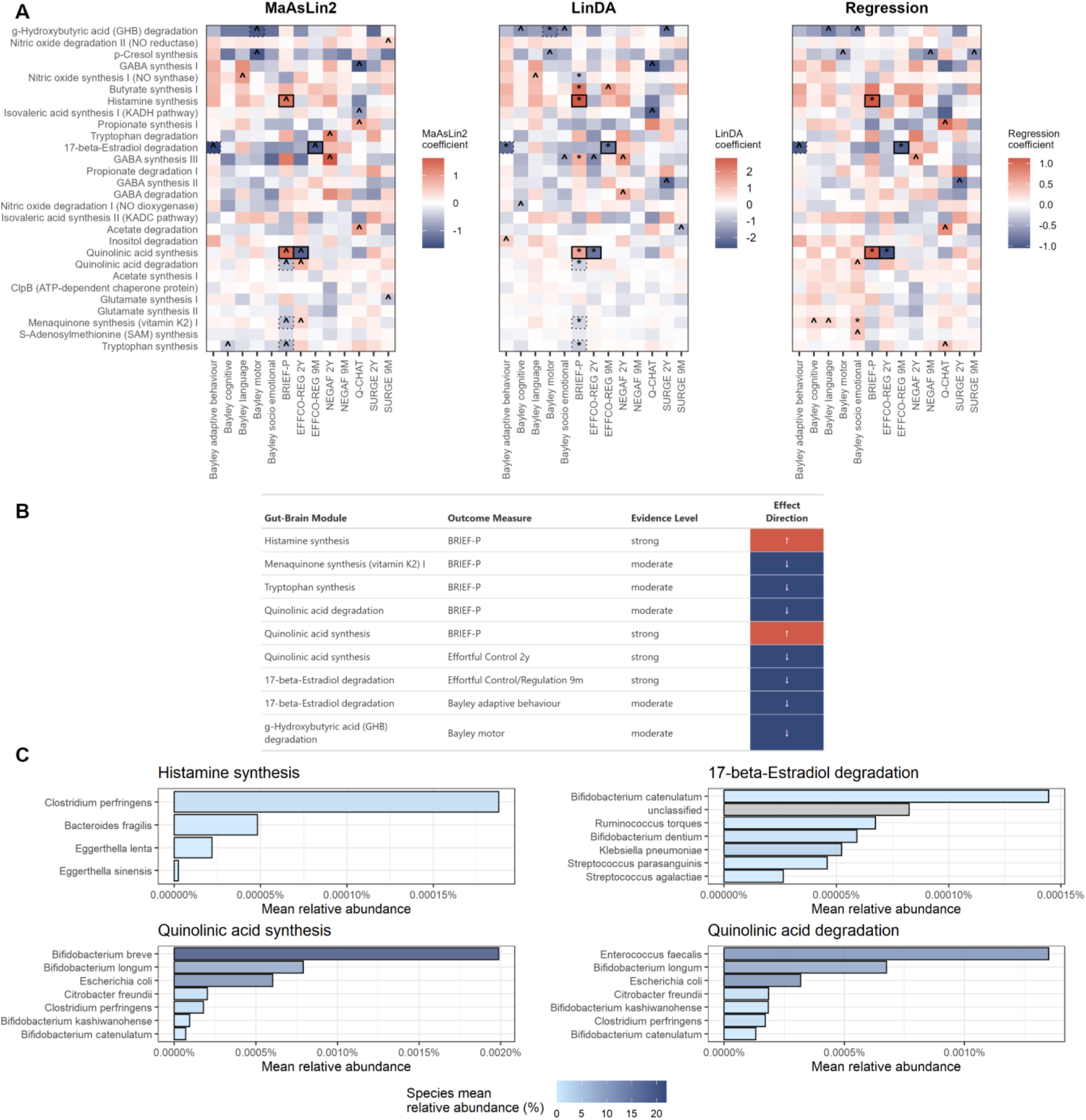
Gut-brain modules associate with executive functioning and adaptive behaviour. (A) Heatmaps of the results of the differential abundance analyses showing associations between gut-brain module abundances and outcome measures of interest. Symbols denote significance level: *statistically significant (q < 0.05 after adjustment for multiple comparisons), ^nominally significant (p < 0.05 prior to adjustment). Boxed cells denote evidence level: solid boxes indicate strong evidence level while dashed boxes indicate moderate evidence level. Modules are ordered based on prevalence, with the most prevalent (i.e. missing in the smallest number of infants in the whole sample) at the bottom. (B) Table highlighting strong and moderate associations observed in the differential abundance analyses. Direction of effect is indicated by colour scheme (blue for negative, red for positive). (C) Relative abundance of the four gut-brain modules associated with neurodevelopmental outcomes stratified by species to show the contributions from known and unknown bacteria; for each module, only the top seven species with the highest abundance of genes present in that module are shown. Depth of the blue colour represents the mean relative abundance of the species in the dataset, with darker colours corresponding to higher overall relative abundance.

Given the observed significant associations between 9-month temperament dimensions and abundances of bacterial species and GBMs, as a sensitivity analysis we repeated the models excluding the 10 infants with imputed scores (see Methods and Supplementary materials). Overall, the pattern of results aligned well with those reported above, both for species (Supplementary figure 4) and GBMs (Supplementary figure 5); the exceptions were the associations for surgency with *Streptococcus parasanguinis* and *Negativicoccus succinicivorans* which were no longer statistically significant. For the three differential abundance methods, the correlations of the model coefficients resulting from the analyses performed with imputed vs non-imputed IBQ data were between 0.87 and 0.97 for species-level and 0.95 and 0.96 for GBM-level analyses, suggesting that the main results were not driven by data imputation.

Regarding the different covariate adjustment strategies, overall, the top findings were similar for the species-level differential abundance analyses in the age-adjusted and fully adjusted models, with some species-outcome associations reducing in magnitude with the inclusion of more covariates as expected (Supplementary tables 8-11). We observed the greatest differences in the top findings for the unadjusted univariable models (Supplementary table 8), with several associations only appearing with the inclusion of age and other covariates. This could be due to noise in the effect estimates given the relatively wide sampling age range. Excluding antibiotic and breast milk covariates (partially adjusted model, Supplementary table 11) also resulted in very similar top results compared to the fully adjusted model, suggesting that the majority of the species-outcome associations are independent of these previously identified early life drivers of microbiota development. However, a few associations changed evidence strength from *moderate* to *strong* (e.g. between *Finegoldia magna* and negative affectivity at 2 years) or from *moderate* to *weak* (e.g. between *K. oxytoca* and negative affectivity at 9 months), or only appeared in one or the other model (e.g. *moderate evidence* for a negative association between *Enterobacter cloacae complex* and Q-CHAT in the partially adjusted model), reflecting that some effects may be sensitive to these covariates. For the GBM-outcome relative abundance analysis, the top findings were also consistent across the unadjusted, age-adjusted and fully adjusted models (Supplementary tables 12-14), though several associations expectedly changed in evidence strength level from stronger to weaker whilst some only appeared as relevant using our evidence level thresholding with the additional adjustment for potential confounders. As with the species-level differential abundance analyses, with the exclusion of antibiotic and breast milk covariates (Supplementary table 15), some associations changed evidence levels between the fully vs partially adjusted models (e.g. a change from *moderate* to *strong* for the negative association between 17-β-estradiol degradation and Bayley adaptive behaviour, and a change from *strong* to *moderate* for the negative association between quinolinic acid synthesis and effortful control at 2 years), while some only appeared in one or the other (e.g. *strong evidence* for an association between quinolinic acid degradation and effortful control at 2 years in the partially adjusted model, and *moderate evidence* for BRIEF-P associations with quinolinic acid degradation, tryptophan synthesis and menaquinone synthesis in the fully adjusted model). In sum, the main results were broadly consistent across the covariate adjustment strategies, with only a minority of associations changing in strength of evidence or appearing uniquely in one model, highlighting the robustness of the main findings and indicating that the inclusion of strong determinants of early life microbiome (breast milk and antibiotic variables) did not result in a substantial over-adjustment.

In previous work^50^, using 16S-based data, there was a good agreement between community-level PCo-based and differential abundance analyses for interpreting the microbiome-brain MRI relationships^50^; however, this agreement was not observed here for neurobehavioral outcomes. Because of this, we conducted an additional supervised data reduction analysis by performing sPLS to identify potential latent components in the microbiome features data that explain variance in the behavioural outcomes and to explore if the species/GBMs contributing to the latent components overlap with those associated with the outcomes in differential abundance analyses. To a large degree, sPLS results were aligned with the main observations from differential abundance testing. For example, the first latent component, which significantly correlated with Q-CHAT scores (r=0.589, explaining 34.7% of the variation) contained all 5 species that showed strong or moderate evidence for association with Q-CHAT in differential abundance analyses; four of these species (all except *S. haemolyticus*) were among the top 5 contributors to this 20-species component (Figure 5A). Similar trends were observed for the Bayley socio-emotional development (Figure 5B) and 2-year negative affectivity measures (Figure 5C), where the species significantly associated with these outcomes in differential abundance analyses, e.g. *Enterobacter cloacae complex* and *Finegoldia magna,* were also selected as the top contributors to the sPLS component. These components explained 28.06% (r=0.530) and 44.9% (r=0.670) of variance in these outcomes, respectively (see Supplementary figure 8 for the loadings of species to the sPLS components and their correlations with the rest of the outcome measures).

**Figure 5.**
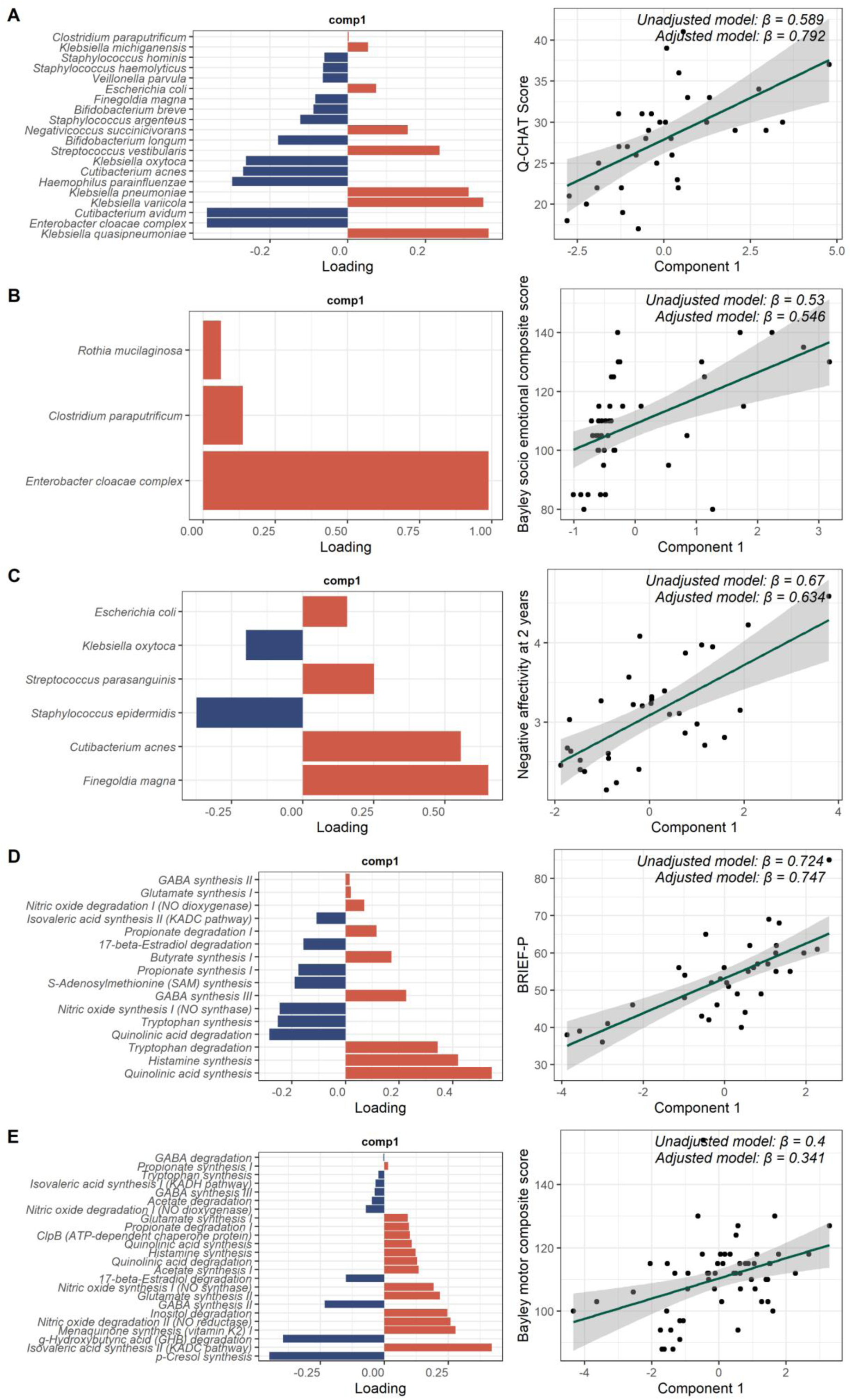
Results of the sparse partial least squares analysis demonstrating the loadings of species or gut-brain modules to the first component (left) and correlation of the first component with the outcome measures (right) for (A) Q-CHAT, (B) Bayley socio-emotional composite score, (C) ECBQ negative affectivity, (D) BRIEF-P and (E) Bayley motor score. Unadjusted β notes the standardised coefficient from the linear regression model associating the sPLS components with the outcome measures, adjusted β notes standardised coefficient from a linear regression model that additionally included GA at birth, PMA at sample, antibiotic exposure < 72h of life, birthweight z-score, high vs low proportion of exclusive breast milk days during NICU stay, infant sex, Scottish Index of Multiple Deprivation quintile, maternal BMI at pregnancy booking, and maternal age as covariates.

The components resulting from sPLS with GBMs also appeared mostly in line with the results of differential abundance testing. For example, 4 out of 5 GBMs correlated with BRIEF-P at strong or moderate evidence level were selected as the contributors to the 16-feature component associated with BRIEF-P with Pearson r=0.724 (Figure 5D). For the other four outcome measures that each had one GBM associated at the strong/moderate evidence level, sPLS similarly selected those GBMs among top 3 strongest contributors to the component associated with those developmental outcomes (Supplementary figure 9).

However, sPLS also selected some other features that were not associated with the outcome measures in the fully adjusted differential abundance analyses, potentially due to the underlying correlational nature of the microbiome data itself. For example, for Q-CHAT, the second greatest contributor to the sPLS component was *Enterobacter cloacae complex* (Figure 5A), which was not identified in the fully adjusted differential abundance analyses, even at the weak evidence threshold, but this association did appear at the *moderate evidence* level in the partially adjusted model. Another example is the latent component identified for 9-month surgency: in addition to the *Veillonella parvula* and *Enterobacter cloacae complex,* which we found significantly associated in differential abundance analyses, the component had additionally high loadings from *Bifidobacterium longum*, *Enterococcus faecalis* and *K. pneumoniae,* among others (Supplementary figure 8E). For GBMs, a contrasting result is that other modules were identified as relatively stronger contributors to the component associated with the Bayley motor domain, such as p-Cresol synthesis and isovaleric acid synthesis (Figure 5E). Conversely, some other diverging results compared to differential abundance testing are noteworthy: in light of the result that tryptophan *synthesis* was an identified module both in differential abundance analyses as well as sPLS in association with BRIEF-P, it is interesting that tryptophan *degradation* was also the third strongest contributor to the BRIEF-P-associated component (Figure 5D), further strengthening the evidence that tryptophan metabolism may be important for infant development.

## 4 Discussion

In a sample of very preterm children we report associations between the neonatal abundances of specific gut bacterial species and functional metabolic modules implicated in gut-brain signalling, and neurodevelopmental outcomes measured during the first two years of life. The associations included autistic traits, executive functioning, socio-emotional development, language skills, and temperament. The specific microbial features associated varied across neurodevelopmental measures, suggesting that gut microbiome–neurodevelopment interactions may be domain-specific and/or temporally dependent. Together, these findings indicate that distinct microbial species and associated metabolic functions may exert selective effects on specific neurodevelopmental pathways. This contributes to a growing body of evidence that the microbiome-gut-brain axis may be important for the neurodevelopment of preterm infants

### 4.1 No association between microbial diversity or community-level variance and neurodevelopmental outcomes

We found limited evidence that general microbiome features, such as alpha and beta diversity, associate with developmental outcomes. Inconsistent results for these measures have been observed previously^18^ and we further add to this literature that microbiome diversity indices may not be adequate markers of gut health or carry prognostic value for future outcomes, especially in early life, although some studies in preterm infants have shown evidence for alpha diversity-outcome relationships^52,54^. Despite observing several associations between overall gut microbiome community composition differences between infants, measured via community composition PCos, and brain MRI features in our previous work^50^, here we do not find evidence that these measures capturing microbiome compositional variance associate with neurodevelopmental outcomes. Instead, it appears that the abundances of specific species and their functional capacity, as will be discussed next, may carry more information for future outcomes. Our supervised data reduction analyses using sPLS further support this, indicating that distinct microbial signatures in early life may be predictive of neurodevelopment, even when general compositional differences are less apparent. In other words, there is limited evidence for a global “dysbiotic state” associated with outcomes, but there may be more specific associations between neonatal gut microbial features and different neurodevelopmental outcomes across infancy and toddlerhood.

### 4.2 Species abundances associate with autistic traits and socio-emotional measures

We found the abundance of several species to be associated with Q-CHAT scores, a screening tool designed to measure autistic traits in toddlers. Both sPLS and differential abundance analyses suggested that higher abundance of multiple *Klebsiella spp*. are positively, and *Cutibacterium avidum* negatively, associated with higher Q-CHAT scores. *Klebsiella spp*. are common colonisers of the preterm infant gut^41,42^, with prior work showing higher abundance of *Klebsiella* in infants exposed to antibiotics during their NICU stay^77–79,50^. Preterm infants are more likely to be diagnosed with autism compared to their term born peers^7^, and indeed preterm toddlers compared to term-born peers show higher Q-CHAT scores in the broader TEBC cohort^80^. *Klebsiella* overgrowth, together with a pro-inflammatory profile in the blood, has been associated with severe brain injury after preterm birth^49^ and we previously demonstrated that several *Klebsiella spp.*, including *K. pneumoniae*, as well as *Cutibacterium avidum*, associated with diffusion MRI microstructural measures of encephalopathy of prematurity in this sample^50^. Interestingly, Q-CHAT-associated *Klebsiella spp*. were top contributors to the GBM for propionate synthesis, a SCFA shown to be associated with ASD^26,81^. Propionate synthesis showed nominal associations with Q-CHAT here and was associated with deep grey matter microstructure in our previous work^50^, providing corroborative evidence that propionate metabolism may mediate gut-brain interactions following preterm birth. We observed different microbiome-autistic trait associations compared to those reported by Laue et al^27^, which could be due to differences in populations (term vs preterm-born), behavioural assessments (Social Responsiveness Scale vs Q-CHAT) as well as timings of microbiome profiling and behavioural measurement. Although high scores on Q-CHAT do not equate to a diagnosis of ASD, and may reflect more general behavioural or socio-emotional differences^66,82^, previous studies have suggested that increased abundance of *Klebsiella* is present in autistic children. A study of 71 autistic boys and 18 neurotypical controls in China found that autistic children had significantly greater relative abundance of *Klebsiella* species compared to controls^83^ and a small study by Abuljadayel et al (2024)^84^ also reported higher abundances of the phylum *Proteobacteria*, including a *Klebsiella* species (*K. oxytoca*) in autistic children compared to their neurotypical siblings. The extent to which our findings can be directly compared to these previous studies is limited, given that we assess early autistic traits using a proxy measure rather than a formal ASD diagnosis, and the microbiome data utilised here were collected in the neonatal period, whereas many prior studies have focused on older children. Nonetheless, these results suggest temporal associations between early-life *Klebsiella* spp. abundance and brain and behavioural outcomes following preterm birth.

From differential abundance testing and sPLS analyses, we also found consistent evidence linking the *Enterobacter cloacae complex* to socio-emotional behaviours: the abundance of this species group was positively associated with the Bayley socio-emotional domain at 2 years and negatively associated with surgency and negative affectivity at 9 months. It also negatively associated with Q-CHAT in sPLS analyses and in differential abundance testing using the model without antibiotic and breast milk exposure covariates, further strengthening the evidence that this species group may be relevant in the context of behavioural outcomes in preterm-born children. *Enterobacter cloacae complex* species are common gut commensals, including in preterm infants^40^, and are frequent causes of hospital-acquired infections, including neonatal sepsis in intensive care settings^85^. To our knowledge, there is little previous evidence directly linking the *Enterobacter cloacae complex* to neurodevelopment. One study reported that, on the genus level, the abundance of *Enterobacter* negatively associated with thalamic volume in two-week-old infants^86^. Moreover, recent research in extremely preterm and extremely low– birth-weight infants found that relatively lower levels of the *Enterobacter* genus, together with higher abundances of *Enterococcus* and *Escherichia*, were observed in neonates who later exhibited poorer neurodevelopmental outcomes at age two^52^. However, as these findings are at the genus level and do not include measures of socio-emotional behaviour, comparisons with the *Enterobacter cloacae complex* specifically are limited. Yet, given we observed several lines of evidence for an association of this group of bacteria with different outcome measures, particularly those related to socio-emotional behaviours, further investigations into the role of *Enterobacter cloacae complex* in modulating the gut-brain axis in preterm infants are warranted.

Moderate evidence was observed for a negative association between *Veillonella parvula* and surgency at 9 months, and weaker evidence for an association with the same temperament domain at 2 years of age. *Veillonella* is a lactate-fermenting bacteria which has been considered a signature taxon in the 4-month-old term infant microbiota^87^ and whose abundance thereafter gradually decreases over the first five years of life^88^ (similar to the trajectory observed for *Bifidobacterium*). While specific associations between *V. parvula* and surgency have not been previously evidenced, associations with other temperament dimensions and affective behaviours have been observed. Aatsinki and colleagues^23^ examined faecal microbiome composition at 6 months and found that, for female infants, higher levels of *V. parvula* and *V. dispar* were associated with lower fear reactivity, and across both sexes, infants with gut microbiota profiles dominated by *V. dispar* had lower surgency scores. On the genus level, a small study of 34 infants found that *Veillonella* was a significant negative driver of a non-social fear related PCo^89^, and another study reported a nominally significant positive association between *Veillonella* abundances at two weeks of life and negative affect at 30 months of age^90^. Furthermore, at two weeks of life, the abundance of *Veillonella* negatively correlated with insula volume^86^. In our previous work in preterm neonates^50^, *V. parvula* also associated with several microstructural indices in the deep grey matter. Collectively, our findings therefore add to the growing body of evidence linking the abundances of *Veillonella* spp. in early life with socio-emotional development, including the structure of brain regions involved in affective and sensory processing in the neonatal period as well as later behavioural manifestations related to affect and temperament. However, further research is required to elucidate the mechanisms through which *Veillonella* spp. may influence brain development, and in turn, infant temperament.

Another noteworthy result is a positive correlation between *Finegoldia magna* and negative affectivity at 2 years of age; this species was also correlated with deep grey matter microstructure in our prior work^50^. *F. magna*, a commensal and opportunistic pathogen which commonly colonises the skin and mucosal surfaces, including the female urogenital tract, has been associated with preterm birth at the genus level^91^, and at the species level with chorioamnionitis^92^ (a common cause of preterm birth). Although, to our knowledge, associations between *F. magna* and negative affectivity have not been reported previously, higher abundances of *F. magna* have been identified in infants of mothers with postpartum depressive symptoms, where it was also negatively associated with fine motor skills^93^. The association observed here between *F. magna* and negative affectivity in preterm infants is novel and highlights a potential avenue for future research.

### 4.3 Gut-brain modules associate with executive functioning, effortful control and regulation

Analyses of the functional potential of the metagenome, both from differential abundance testing as well as sPLS models, showed strong evidence for associations between several gut-brain modules, including histamine synthesis and modules related to quinolinic acid and tryptophan metabolism, and executive functioning (EF) difficulties measured with BRIEF-P. EF, including working memory, cognitive flexibility, attention and inhibitory control, are frequently affected in children born preterm^94–96^, even after controlling for IQ^97^. These differences are important, given EF abilities have been consistently linked to academic success across childhood, adolescence, and adulthood^98–101^. Lower EF scores, including assessment using the BRIEF instrument which we used here, have also been associated with later risk for psychopathology, including depression, anxiety, attention-deficit/hyperactivity disorder (ADHD), and obsessive-compulsive disorder, as well as with alterations in resting-state functional brain connectivity^102–104^. This suggests that neonatal microbiome, through modulating EF in toddlerhood, may have wide-ranging implications for preterm children. To our knowledge, only one study has specifically assessed correlations between gut microbiota composition and EF in childhood, finding no evidence for associations between genus level bacterial relative abundances and EF^105^. Our use of shotgun sequencing, which allowed probing of the functional capacity, and focus on the high-risk preterm population, may have allowed us to detect previously uncovered relationships.

Histamine, a neurotransmitter with several nervous system functions including synaptic plasticity, neurotransmission and modulation of the immune system^106,107^, has been associated with attention, wakefulness, memory and cognition^108,109^, and peripherally (including gut) derived histamine can impact the brain indirectly, particularly under conditions of inflammation^75,76^. Histamine also plays an important role in brain development^110^, with early developing histamine immunoreactive cells observed in the human foetal brain at 19 weeks and more mature-like histaminergic neurons observed by 36 weeks gestation^111^, indicating that the histaminergic system develops progressively throughout mid-to-late gestation. This suggests that in very early or extreme preterm birth, histamine reactive cells and circuits may be mid-development at the time of birth. Given this immaturity, the developing brain may be more vulnerable to increased peripheral histamine production, particularly in the context of increased inflammation.

Interestingly, *Clostridium perfringens*, the species which was the strongest contributor to the histamine synthesis GBM, and which was positively associated with BRIEF-P, has been highlighted previously for its ability to produce high concentrations of histamine *in vitro*^112^. In addition, it was more frequently identified in patients with histamine intolerance (a result of histamine build-up in the body) vs healthy individuals^113,114^. Together, these findings suggest that *Clostridium perfringens* may contribute to increased activation of microbial histamine synthesis pathways, and this may represent one mechanism contributing to differences in cognitive outcomes, in this case, BRIEF-P scores.

Relationships were also observed between quinolinic acid metabolism and EF difficulties in multiple measures including BRIEF-P and ECBQ effortful control domain. Quinolinic acid is an excitotoxic NMDA receptor agonist and a downstream metabolic product of tryptophan degradation through the kynurenine pathway^115^. Elevated levels of quinolinic acid have been associated with several neurocognitive and neurodegenerative conditions including Alzheimer’s disease^116,117^, ASD^118^ and schizophrenia^119^, while imbalances in the ratio of kynurenic acid to quinolinic acid have been observed in mood disorders^120^. sPLS analysis confirmed the differential abundance findings, with tryptophan and quinolinic acid metabolism among the top GBMs contributing to the BRIEF-P- and effortful-control-associated components. Interestingly, we previously showed that tryptophan degradation GBM associated with deep grey matter microstructure^50^. These observations further support the relevance of tryptophan-related metabolism, particularly the kynurenine pathway, to neurodevelopmental outcomes involving EF, as suggested previously^121^.

As with histamine synthesis, *Clostridium perfringens* was among the top 7 species with the highest abundance of genes involved in quinolinic acid synthesis and degradation. Although seemingly contradictory, it is not unusual for microbial species to carry genes for both the synthesis and degradation of a given compound, with functional expression depending on multiple factors, including environmental conditions. For example, a murine clostridial myonecrosis infection model showed that under inflammation-like conditions, *Clostridium perfringens* exhibits a significantly altered gene expression profile^122^, highlighting how environmental cues may potentially influence which metabolic pathways are favoured. Indeed, we also found several *Bifidobacterium spp*. among the top contributors to these quinolinic acid metabolism pathways, yet the abundance of these *Bifidobacterium spp*. themselves was not significantly correlated with outcomes, further highlighting the differences between the abundance of a certain bacterial species, the functional capacity of the microbes, and the expression of this function. Future studies with metatranscriptomic or targeted metabolomics analyses are needed to clarify the role of quinolinic acid metabolism in neurodevelopment, and the bacteria involved in this process.

BRIEF-P also showed a negative association with menaquinone (Vitamin K2) synthesis in the fully adjusted model. Vitamin K2 is synthesised by gut bacteria^123,124^. Supplementation with Vitamin K2 has been proposed as a potential intervention to mitigate cognitive decline associated with Alzheimer’s disease^125,126^ and it has shown cognitive benefits in rodent models. For example, a study in neonatal mice exposed to repeated anaesthesia—which induced “ADHD-like” behaviours such as impulsivity, hyperactivity, and cognitive impairment—found that administering Vitamin K2 30 minutes prior to each anaesthetic procedure significantly reduced these cognitive deficits^127^. Interestingly, several *Klebsiella* spp., including *K. variicola* and *K. oxytoca*, which showed weak levels of evidence for associations with BRIEF-P and effortful control, respectively, were among the top contributors to this functional module. Our results therefore add to the literature supporting the role of Vitamin K2 in neural functions and highlight the need of future studies investigating gut microbiome-produced Vitamin K2 in neurodevelopment.

This study is among the first to provide evidence that neonatal gut microbiome, and especially it’s functional capacity, may correlate with EF and effortful control in 2-year-old preterm toddlers, motivating future studies to investigate this question in more detail. To reduce the number of tests performed in our relatively small sample, we only used the global executive composite of BRIEF-P. Yet, this rating scale consists of multiple subdomains targeting different aspects of EF, such as inhibitory control and impulsivity, task shifting ability, emotion regulation/control, planning, and working memory. The ECBQ effortful control dimension similarly captures a range of behaviours related to inhibitory control, attentional focussing and shifting. Future work should clarify the specific subdomains of EF most strongly associated with variation in early life microbiome.

Beyond BRIEF-P and effortful control, the abundance of 17β-estradiol degradation GBM negatively associated with the temperament domain of regulation at 9 months of age, and the Bayley adaptive behaviour domain at 2 years. Additionally, it was amongst the top component contributors for both of these outcome measures in sPLS analysis. 17β-estradiol is a steroid hormone and the primary form of oestrogen in humans. 17β-estradiol, produced by the placenta^128–130^, plays a key role in foetal neurodevelopment, including sexual differentiation of the brain, synaptogenesis, myelination, and neuroprotection^131–133^. Several animal studies have demonstrated that estradiol treatment can reverse or mitigate the neurodevelopmental disruptions caused by prematurity, supporting neurogenesis, synapse formation, and cognitive outcomes^134–136^, though clinical evidence remains limited^137^. Regarding the metabolism of 17β-estradiol by the gut microbes, a recent study found that the gut microbiota of premenopausal women with depression degraded nearly 80% of serum 17β-estradiol within two hours, compared to just 19% degradation by microbiota from age-matched, non-depressed controls^138^. The researchers identified specific oestradiol-degrading bacteria, such as *Klebsiella aerogenes*, and demonstrated that transplantation of these microbes into recipient mice led to reduced serum estradiol levels and the onset of depressive-like behaviours. Interestingly, in our work, *Klebsiella pneumoniae* was among the top species with the highest abundance of genes for 17β-estradiol degradation. Although this species was not significantly associated with either regulation at 9 months or Bayley adaptive behaviour, it associated with higher Q-CHAT scores. The association between 17β-estradiol and regulation and adaptive behaviour observed in this work is novel and may warrant further investigation, particularly in light of the well-documented sex-related differences in neurodevelopmental outcomes amongst preterm infants^139,140^.

Finally, it is important to highlight that for some outcomes which have previously been associated with gut microbiome features, we did not observe significant associations. For example, motor skills were not associated with any species’ abundances, and only weak to moderate evidence for associations with the abundances of two GBMs was observed. Furthermore, the Bayley cognitive domain only showed weak evidence for an association with the abundances of a single bacterium. This could result from several methodological differences such as the study of exclusively preterm-born individuals, the specific timepoint selected for microbiome profiling, use of shotgun rather than 16S-based sequencing and our consensus-based statistical analyses.

### 4.4 Cross-method concordance strengthens confidence in differential abundance results

The high dimensionality of gut microbiome data, combined with substantial analytical flexibility available, has likely contributed to inconsistent findings in the literature. Therefore, in this preregistered work we opted for a consensus-based approach. We employed multiple data modelling techniques, including three differential abundance testing methods with distinct data transformations and underlying assumptions alongside sPLS analysis, and focussed on results that converge across the methods. Overall, we observed strong concordance across methods, which strengthens confidence in the robustness of our findings. However, the slight variation in statistically significant results across individual differential abundance testing methods suggests that relying on a single approach could yield divergent conclusions. This underscores the value of using a consensus approach in microbiome analyses, as has been previously suggested^141^.

### 4.5 Strengths and limitations

This study has several strengths. Neonatal gut microbiome linked to longitudinal behavioural data in very preterm infants, who have a high likelihood for atypical neurodevelopment, is a rare resource. Shotgun metagenomic sequencing allowed us to comprehensively characterise the associations both for the taxonomic composition at the species level and as well as the functional potential of the metagenome. The latter was profiled by specifically focussing on the GBMs – curated groups of functional processes involved in gut-brain signalling – which allowed for a principled approach to data reduction. We included a wide battery of assessments, incorporating both parental reports as well as direct observational measures, to assess development comprehensively. We adopted multiple covariate adjustment strategies and were able to show that the main findings were broadly consistent across models, highlighting the robustness of the main findings. Lastly, the consensus-based approach adopted here provides a particular strength to this work, adding robustness and confidence in the reported results.

Several limitations of the current study were also identified. The relatively small sample size for microbe-outcome associations, partially due to some loss of follow-up from the neonatal period to toddlerhood during the pandemic, may have limited statistical power to detect smaller effect sizes and precluded detailed analyses of sex- or GA-specific microbiome–outcome associations. Next, whilst the microbiome sampling timepoint was chosen to reflect the cumulative effects of preterm birth and NICU exposures, the single timepoint design limited our ability to examine whether specific developmental windows are particularly sensitive to microbiome influences; future studies with longitudinal sampling over the course of NICU stay are needed to further clarify the critical windows of microbial influence on neurodevelopment. Additionally, although the cohort is representative of most survivors of preterm birth in high income settings in terms of clinical features and brain MRI findings^142^, the single-centre nature of recruitment may impact the degree to which these findings can be generalised to infants cared for in other neonatal units where practices may differ. Future studies are also encouraged to include a term comparator group in microbiome-outcome investigations to determine whether the observed associations are specific to preterm infants or also apply to those born at term, and to explore if microbiome compositions more closely resembling those of term infants are associated with improved neurodevelopmental outcomes following preterm birth. Given the observational nature of this study, causal inferences cannot be drawn as the relationships observed could arise due to other shared processes affecting both the gut microbiome and behavioural development.

## 5 Conclusion

This study identified specific features of the neonatal gut microbiome associated with neurodevelopmental outcomes during the first 2 years of life in preterm-born children. These include bacterial species linked to autistic traits, socio-emotional development, and temperament. Notably, novel links were observed between microbial functional potential and early EF in this population. Distinct associations between microbiome features and outcome domains suggest that microbiome– neurodevelopment relationships may be domain-specific, and the use of multiple analytical approaches strengthened the robustness of findings. Collectively, these results contribute to growing evidence that early gut microbiome composition and functional capacity may influence later neurodevelopment. Our findings may carry importance for the development of future microbiome-based risk stratification tools or microbiome-modification based therapies, including probiotics, to support development following preterm birth, and highlight the need to incorporate longitudinal neurodevelopmental assessments, including those related to autism, socio-emotional development, and executive functioning, into future studies evaluating such tools and therapies in preterm infants.

## Supporting information

Supplementary table 1 (STORMS checklist)

## 6 Acknowledgments

This research was funded in part by the Wellcome (no. 108890/Z/15/Z, 228326/Z/23/Z, 218493/Z/19/Z). For the purpose of open access, the author has applied a CC BY public copyright license to any Author Accepted Manuscript version arising from this submission. This work was supported by Theirworld (www.theirworld.org). K.V., A.K., L.J-S. and R.S. were supported by the Translational Neuroscience PhD Program at the University of Edinburgh, funded by Wellcome (grant numbers 108890/Z/15/Z, 228326/Z/23/Z and 218493/Z/19/Z). J.P.B. is supported by a UKRI MRC programme grant (MR/X003434/1). H.T. is a PhD student in Clinical Brain Sciences and receives funding support from Simpson Special Care Babies charity. Collection of Bayley-III data was funded by NHS Lothian. The authors are grateful to the families who consented to take part in the study. We acknowledge Heleen de Weerd and Edinburgh Genomics for executing the bioinformatic processing of the shotgun data; shotgun metagenomic sequencing was performed at Novogene. ChatGPT-5 was utilised as an aid to improve conciseness, clarity and flow while writing this manuscript. Its use was restricted to linguistic editing, and the lead authors thoroughly reviewed, adapted and edited the outputs where appropriate. The authors take full responsibility for the final text.

## 7 Author contributions

Conceptualisation: K.V., A.K., D.B., J.P.B.

Methodology: K.V., A.K., D.B., J.P.B.

Investigation: A.C. (participant recruitment, sample collection and storage); A.C., K.V. (clinical data collection); K.V., P.L.P. (laboratory work, execution and quality control); K.V., L.J-S., H.T., R.S., A.C., H.C., M.R., S.F.-W. (neurobehavioural outcome data collection and quality checking)

Data Curation: K.V. Formal Analysis: K.V., A.K. Visualisation: K.V., A.K.

Writing – Original Draft: K.V., A.K.

Writing – Review & Editing: D.B., J.P.B., all authors Supervision: D.B., J.P.B.

Funding Acquisition: K.V., A.K., D.B., J.P.B

All authors contributed to the design of the analyses prior to preregistration, interpretation of results, critical revision of the manuscript, and approved the final version.

