## Supplementary materials and figures for "From microbes to milestones: Gut bacterial abundances and functional pathways associate with neurodevelopment following preterm birth"

#### Supplementary methods

##### Antibiotic treatment regimen

The antibiotic treatment for all preterm infants with suspected and confirmed neonatal sepsis conformed to the following principles: babies up to 72 h of age were commenced on benzylpenicillin and gentamicin, babies >72 h of age were commenced on piperacillin/tazobactam and vancomycin. To reduce unnecessary exposure to antibiotics, treatment was stopped after 48 h if blood cultures were negative and the clinician had a low suspicion about infection. Some infants in the cohort were also treated with azithromycin, cefotaxime, co-amoxiclav, flucloxacillin, linezolid, meropenem and metronidazole according to symptoms and diagnostic results.

Antibiotic exposure was assessed by three composite variables: (i) exposure to antibiotics during the first three days of life, (ii) exposure to antibiotics at any other time during the NNU stay, and (iii) proportion (%) of antibiotic exposure days during NNU stay (total number of antibiotic treatment days was divided by the number of days in NNU) (see Supplementary table 2). No infants were excluded based on antibiotic exposure.

##### Gut microbiome methods detail

***Faecal sample collection and processing.*** Faecal material was collected from dirty diapers by parents, NICU staff or research team. Samples for microbiome sampling were obtained from preterm infants shortly prior to their discharge from the NICU (for details see our previous work[1]), which was around term-equivalent age. The samples were frozen at -20°C directly after sample collection prior to transfer on ice to a -80°C freezer in the Queens Medical Research Institute (QMRI, University of Edinburgh) until proceeding with DNA isolation; no preservation buffers were used.

***DNA isolation.*** Bacterial DNA was extracted as previously described using a protocol that involves phenol/bead beating in combination with the Mag Mini DNA Isolation Kit (LGC genomics, Germany), previously known as '*Agowa* Mag Mini DNA Isolation Kit'[1,2]. Samples were thawed on ice for as little time as possible to obtain one 10 µl inoculation loop of raw faeces which was added to a 2 ml screwcap tubes containing a mixture of 150 µl lysis buffer (Mag Mini DNA Isolation Kit, LGC genomics, Germany), 0.1 mm zirconium beads (BioSpec products, USA) in 650 µl lysis buffer, and 500 µl of phenol saturated with Tris-HCl (pH 8.0; BioSpec products, USA). The samples were mechanically disrupted twice for 2 minutes at 2100 oscillations/minute using a bead beater (BioSpec products, USA). The samples were then centrifuged for 10 minutes at 5000 rpm at room temperature. Then, the aqueous phase was added to 1300 µl of binding buffer (Mag Mini DNA Isolation Kit) with 10 µl magnetic beads (LGC genomics, Germany) in a sterile 1.5 ml Eppendorf tube and incubated for 30 minutes at room temperature on a thermos shaker (Hettich lab technologies, USA) to allow DNA binding. Subsequently, the supernatant was discarded. The magnetic beads were washed twice with wash buffer 1 (Mag Mini DNA Isolation Kit,), once with wash buffer 2 (Mag Mini DNA Isolation Kit), and air-dried for 15 minutes at 55°C. DNA was eluted in 50 µl elution buffer. DNA isolation from the pre-NICU discharge samples was performed in five batches in a span of a few weeks. Each extraction batch was accompanied by negative (200 µl of lysis buffer) and positive controls (ZymoBIOMICS Microbial Community Standard [Zymo Research, USA] and/or a convenience saliva sample). The amount of extracted bacterial DNA was determined by quantitative polymerase chain reaction (qPCR) as previously[2,3] with universal primers and probes targeting the 16S-rRNA gene (forward: 5'-CGAAAGCGTGGGGAGCAAA-3', reverse: 5'-

GTTCGTACTCCCCAGGCGG-3', TAMRA probe: 6FAM-ATTAGATACCCTGGTAGTCCA-MGB; Life Technologies, USA).

***Metagenomic shotgun sequencing and bioinformatic processing.*** Samples with 16S qPCR concentration >0.8ng/µl (97 samples from pre-NICU discharge, mean [range] 16S qPCR concentration 13.40 [0.85 – 46.87] ng/µl) alongside saliva positive controls and isolation negative controls were considered for shotgun metagenomic sequencing at Novogene facility (Novogene Co., Ltd, Cambridge, UK). Sequencing was performed on the NovaSeq 6000 platform (Illumina) with a read length of 150-bp paired-end reads producing 9G raw data per sample. Shotgun sequencing failed for all negative controls, indicating absence or very low abundance of biological material, illustrated also by the very low 16S qPCR concentration in isolation negative controls ($3.94 \times 10^{-5}$ [$9.32 \times 10^{-6}$ – $1.16 \times 10^{-4}$] ng/µl).

Data pre-processing and annotation was performed by Edinburgh Genomics. Across all samples included in our previous work[1], whole metagenome shotgun sequencing produced a mean of 37410649 (range 2512236 – 81101422) raw reads per sample. The raw reads were cleaned using cutadapt (v3.5)[4]. Adapters were removed, reads were cut when the quality dropped below 30, and reads shorter than 50 bases were removed. Reads belonging to the host were removed by bowtie2 (v2.4.1)[5,6] using Homo Sapiens (GRCh38) as a reference. Files sequenced on multiple lanes but belonging to the same sample were merged into single forward and reverse files.

Taxonomic profiling was performed using MetaPhlAn (v3.1)[7] with the standard database (mpa_v31_CHOCOPhlAn_201901). Functional profiling was performed using HUMAnN (v3)[7] with the default chocophlan (chocophlan.v201901_v31) and the uniref90 databases (uniref90_annotated_v201901b). As MetaPhlAn and HUMAnN do not use paired information of reads, all reads of a sample were merged into a single file and used for taxonomical/functional assignment. As shotgun metagenomic sequencing was performed on higher density samples, the relative contribution of potentially contaminant taxa is smaller, thus, no further quality controls/decontamination on species/functional level were performed. Functional gene family data in reads per kilobase (RPK) units was grouped to KEGG orthogroups (KO-s) using the *humann_regroup_table* function, both for community-level totals and species-stratified gene families. The KO abundance data table was further total-sum-scaled (TSS) to relative abundances using the *humann_renorm_table* function; the unmapped and ungrouped reads were taken into account for TSS-normalisation but excluded from downstream statistical analyses. We computed gut-brain modules (GBM)[8] from both the normalised KO table as well as the KO table in RPK units using the *omixer-rpmR* library[9] and the default settings for both community and taxon-stratified levels.

***Sample inclusion.*** As detailed in our previous publication[1], a total of 104 preterm infants provided a stool sample prior to NICU discharge; one infant was excluded due to congenital abnormality. Out of the 103 remaining infants, 97 had samples with sufficient DNA yield (>0.8 ng/µl) for shotgun metagenomic sequencing. We excluded two additional infants because their samples were collected at a very early postmenstrual age (<33 weeks) prior to infant transfer to another neonatal unit. Of the 95 infants, 73 had data available for any of the developmental outcome measures of interest.

#### Infant Behaviour Questionnaire (IBQ) and Early Childhood Behavioural Questionnaire (ECBQ)

IBQ/ECBQ was administered at three timepoints in Theirworld Edinburgh Birth Cohort (TEBC): at 4.5 and 9 months, and at 2 years. From the questionnaire data, we calculated three broad dimensions of temperament: Negative Affectivity, Surgency/Extraversion, and Regulation (equivalent to Effortful Control domain in ECBQ). We found that the scores at 4.5 and 9 months were strongly correlated (Pearson correlation coefficients between 0.55 to 0.58, n=150 across the entire TEBC dataset), therefore, to reduce number of tests, 4.5-month IBQ data was not included in this work. Instead, with the aim of increasing sample size, we used the entire TEBC dataset with matching 4.5 and 9 month IBQ data to predict 9 month temperament scores for those infants with only 4.5 month data. For each of the three temperament dimensions separately, we built a linear regression model predicting 9 month scores from 4.5 month scores, GA at birth, corrected age at 4.5 month appointment, and infant sex. The mean absolute errors between predicted and actual 9 month scores were 0.44, 0.43 and 0.52 for surgency, regulation and negativity, respectively. These errors were considerably smaller than the standard deviation of the measures (0.69, 0.67 and 0.77 for surgency, regulation and negativity, respectively). We then used these models to predict 9 month scores for those infants with only 4.5 month data. Visualisation of the data indicated that the scores for infants with predicted data overlapped well with those with actual data (i.e. the infants with predicted scores did not have systematically lower/higher scores than those with actual data; not shown). Of infants with available microbiome data, we could use this approach to predict 9 month IBQ dimension scores for 10 infants. One infant's data was excluded from 2-year ECBQ due to a substantial number of original questionnaire items unanswered.

#### Covariate identification

We considered the following variables as potential covariates given their previously demonstrated potent influence on early life microbiome: GA at birth, birthweight z-score, infant sex, delivery mode, antibiotic exposure (<72h or >72h of life), proportion of inpatient days with antibiotic treatment, high vs low proportion of exclusive breastfeeding days during NICU stay, diagnosis of BPD, and sepsis. Necrotising enterocolitis (NEC) was not considered because infants with history of NEC did not have matching outcome data available; birthweight was also not considered because of high collinearity with GA at birth (Spearman $\rho$ = 0.795). We additionally considered the following variables given their relationships with neurodevelopmental outcomes: maternal age[10], maternal BMI[11], maternal education[12], maternal postnatal depression score[13], and socioeconomic status[14,15]. These have also been shown to modify offspring microbiome in previous studies[16–20]. Age at microbiome sampling (postnatal or postmenstrual) does not meet the definition of confounder, because it cannot be causally linked to outcome measures in infancy but was adjusted for here to increase precision in model estimates. In alignment with our previous study[1], we adjusted for postmenstrual age (PMA) rather than postnatal age at sampling due to the high collinearity between GA at birth and postnatal age at sample collection (Spearman $\rho$ = -0.890), which restricts the inclusion of both in statistical models. Description and coding of these covariates is described in Supplementary Table 1.

#### Supplementary tables

Supplementary Tables 1 (STORMS), and 4-15 are provided in separate Word and Excel files, respectively.

***Supplementary Table 2. Description and coding of potential covariates. Those included in the final fully adjusted model are indicated by blue shading.***

| Variable | Description and coding |
|---|---|
| Infant sex | Male or female |
| Infant gestational age at birth | Weeks |
| Birthweight z-score | Weight z-score calculated based on the International Fetal and Newborn Growth Consortium for the 21st Century (INTERGROWTH-21st) standards for preterm infants [21] |
| Infant gestational/postmenstrual age at faecal sample collection | Weeks |
| Delivery mode | C-section (elective or emergency) or vaginal delivery (spontaneous or assisted) |
| Antibiotic exposure <72h of life | Yes or no |
| Antibiotic exposure >72h of life | Yes or no |
| Proportion of antibiotic exposed days during NICU stay | Total number of days of antibiotic treatment received divided by the number of days in NICU |
| Infant exclusive breast milk exposure during NICU stay | Each day in NICU was categorised as consisting of exclusive maternal breast milk feeds, exclusive formula milk feeds, exclusive donor expressed milk feeds, or any combination of these feeding types. Data was available as the sum of each milk type over the entire duration of NICU stay. Breast milk exposure was defined as the proportion of inpatient days infants received exclusive breast milk feeds, which included both maternal and/or donor expressed breast milk.<br>Infants were categorised into two groups based on breast milk exposure: high breast milk exposure was defined as exclusive breast milk feeds for ≥ 75% of inpatient days and low breast milk exposure was defined as exclusive breast milk feeds for < 75% of inpatient days[22,23]. |
| Bronchopulmonary dysplasia | Defined as the requirement for supplemental oxygen or respiratory support at 36 weeks gestational age |
| Sepsis | Defined as detection of a bacterial pathogen from blood culture, or physician decision to treat with antibiotics for ≥ 5 days in the context of growth of coagulase negative *Staphylococcus* from blood or a negative culture but raised inflammatory markers in blood. |
| Maternal age | Years |
| Maternal BMI at pregnancy booking | kg/m2 |
| Maternal education (i.e. mother's final educational qualification) | Data was obtained as following:<br>1 = none<br>2 = 1-4 National 5s / Standard Grades / General Certificate of Secondary Education<br>3 = > 5 National 5s / Standard Grades / General Certificate of Secondary Education<br>4 = A levels / Highers / equivalent<br>5 = College qualification (e.g. National Certificate, Higher National Certificate, Higher National Diploma)<br>6 = University undergraduate degree<br>7 = University postgraduate degree<br>From this data we created a dichotomous variable by combining brackets 1-5 and 6-7 to indicate whether the mother had obtained a university/postgraduate degree. |
| Maternal postnatal depression score | Score of 10 or higher on the self-reported Edinburgh Postnatal Depression Scale[24] at term-equivalent age. |
| Scottish index for multiple deprivation (SIMD) quintile | Derived from the family's postcode at time of birth[25] |

*Supplementary Table 3. Primary software and packages used in this work.*

| Software or package | Source/reference | Link |
|---|---|---|
| Analysis code and scripts | This paper | https://git.ecdf.ed.ac.uk/jbrl/microbiome-and-developmental-outcomes |
| cutadapt v3.5 | Martin 2011[4] | https://github.com/marcelm/cutadapt/tree/9276a89d5df9282ff51602b042b40c3b48c98566 |
| bowtie2 v2.4.1 | Langmead et al 2019[5], 2012[6] | https://bowtie-bio.sourceforge.net/bowtie2/manual.shtml |
| MetaPhlAn v3.1 | Beghini et al.[7] | https://github.com/biobakery/MetaPhlAn |
| HUMAnN v3 | Beghini et al.[7] | https://github.com/biobakery/humann |
| R v4.4.2, v4.5.0 (Maaslin2 and sPLS) | R core team | https://www.r-project.org |
| tidyverse v2.0.0 | Wickham et al[26] | https://tidyverse.tidyverse.org/index.html |
| here v1.0.1 | Müller[27] | https://here.r-lib.org/ |
| phyloseq v1.50.0, v1.52.0 | McMurdie and Holmes[28] | https://joey711.github.io/phyloseq/ |
| vegan v2.6-10, v2.7-1 | Oksanen et al.[29] | https://cran.r-project.org/web/packages/vegan/index.html |
| omixerRpm v0.3.3 | Darzi et al.[9]; Vieira-Silva et al.[30]; Valles-Colomer et al.[31] | http://www.raeslab.org/gomixer/ |
| ape v5.8-1 | Paradis & Schliep[32] | https://cran.r-project.org/web/packages/ape/index.html |
| ppcor v1.1 | Kim[33] | https://cran.r-project.org/web/packages/ppcor/index.html |
| Maaslin2 v1.22.0 | Mallick et al.[34] | https://bioconductor.org/packages/release/bioc/html/Maaslin2.html |
| Tjazi v0.1.0.0 | Bastiaanssen et al.[35] | https://github.com/thomazbastiaanssen/Tjazi |
| MicrobiomeStat v1.2 (LinDA) | Zhou et al[36] | https://cran.r-project.org/web/packages/MicrobiomeStat/index.html |
| ggplot2 v3.5.2 | Wickham[37] | https://ggplot2.tidyverse.org/ |
| ggpubr v0.6.0 | Kassambara[38] | https://rpkgs.datanovia.com/ggpubr/ |
| cowplot v1.1.3 | Wilke[39] | https://wilkelab.org/cowplot/ |
| gridExtra v2.3 | Auguie[40] | https://cran.r-project.org/web/packages/gridExtra/index.html |
| Hmisc v5.2-3 | Harrell[41] | https://cran.r-project.org/web/packages/Hmisc/index.html |
| mixOmics v6.32.0 | Rohart et al[42] | https://mixomics.org/ |
| RColorBrewer v1.1-3 | Neuwirth[43] | https://cran.r-project.org/web/packages/RColorBrewer/index.html |
| gtsummary v2.2.0 | Sjoberg et al[44] | https://doi.org/10.32614/RJ-2021-053 |
| corrplot v0.95 | Wei & Simko[45] | https://taiyun.github.io/corrplot/ |
| effsize v0.8.1 | Torchiano[46] | https://cran.r-project.org/web/packages/effsize/index.html |
| ggstatsplot v0.13.1 | Patil[47] | https://cran.r-project.org/web/packages/ggstatsplot/index.html |
| patchwork v1.3.0 | Pedersen[48] | https://patchwork.data-imaginist.com |
| scales v1.4.0 | Wickham et al[49] | https://scales.r-lib.org |
| ggdendro v0.2.0 | de Vries & Ripley[50] | https://andrie.github.io/ggdendro/ |
| magick v2.8.7 | Ooms[51] | https://CRAN.R-project.org/package=magick |
| gt v1.0.0 | Iannone et al[52] | https://gt.rstudio.com |

### Supplementary Figures

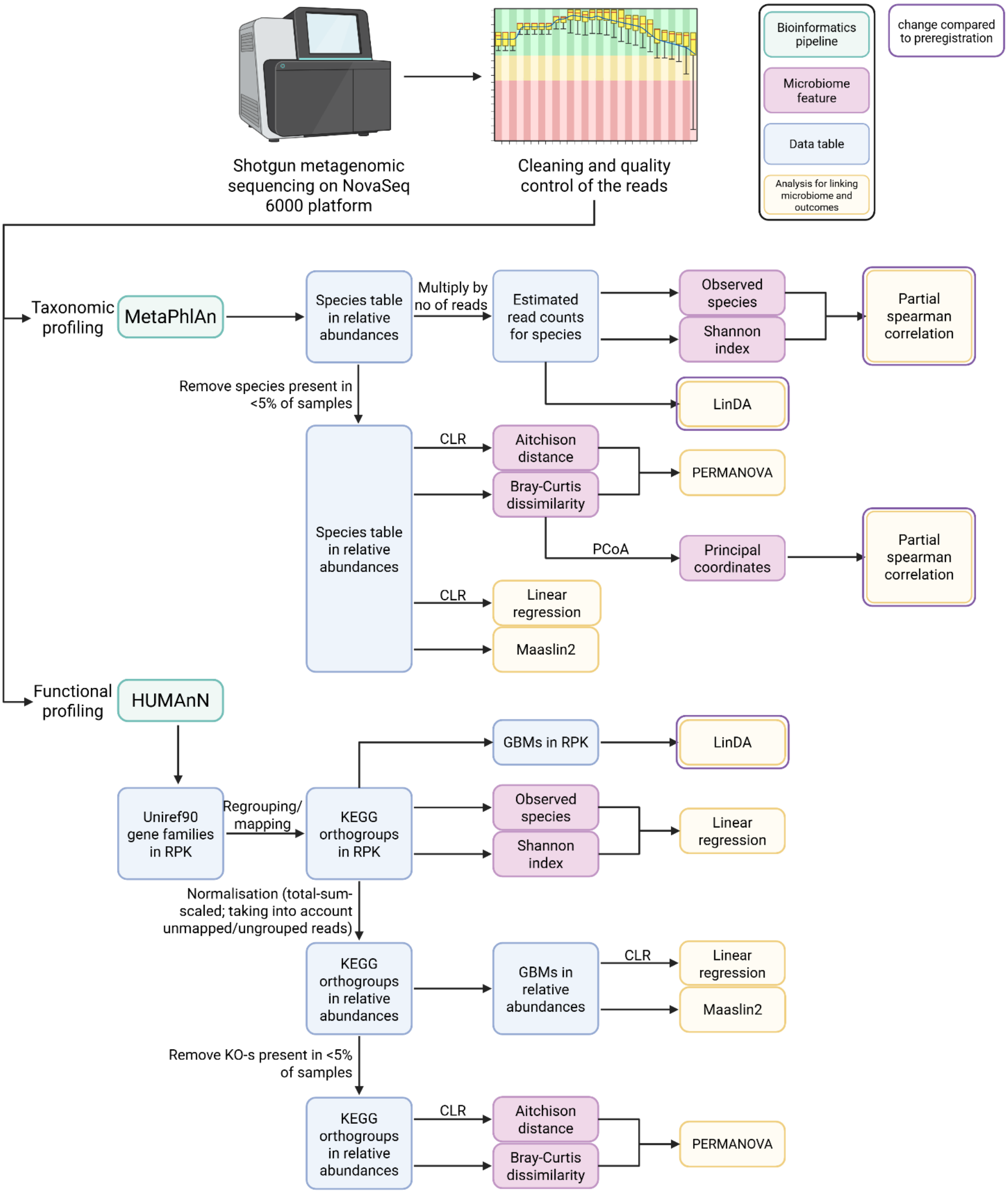


***Supplementary Figure 1. Scheme for a statistical analysis linking preterm infant microbiome data with neurodevelopmental outcomes. See main text for details. Created with BioRender. CLR = centered log ratio, PERMANOVA = permutational analysis of variance, GBM = gut-brain module, PCoA = principal coordinates analysis, Maaslin = Microbiome Multivariable Association with Linear Models, LinDA = linear regression framework for differential abundance analysis, KEGG = Kyoto Encyclopedia of Genes and Genomes, KO = KEGG orthogroup, RPK = reads per kilobase.***

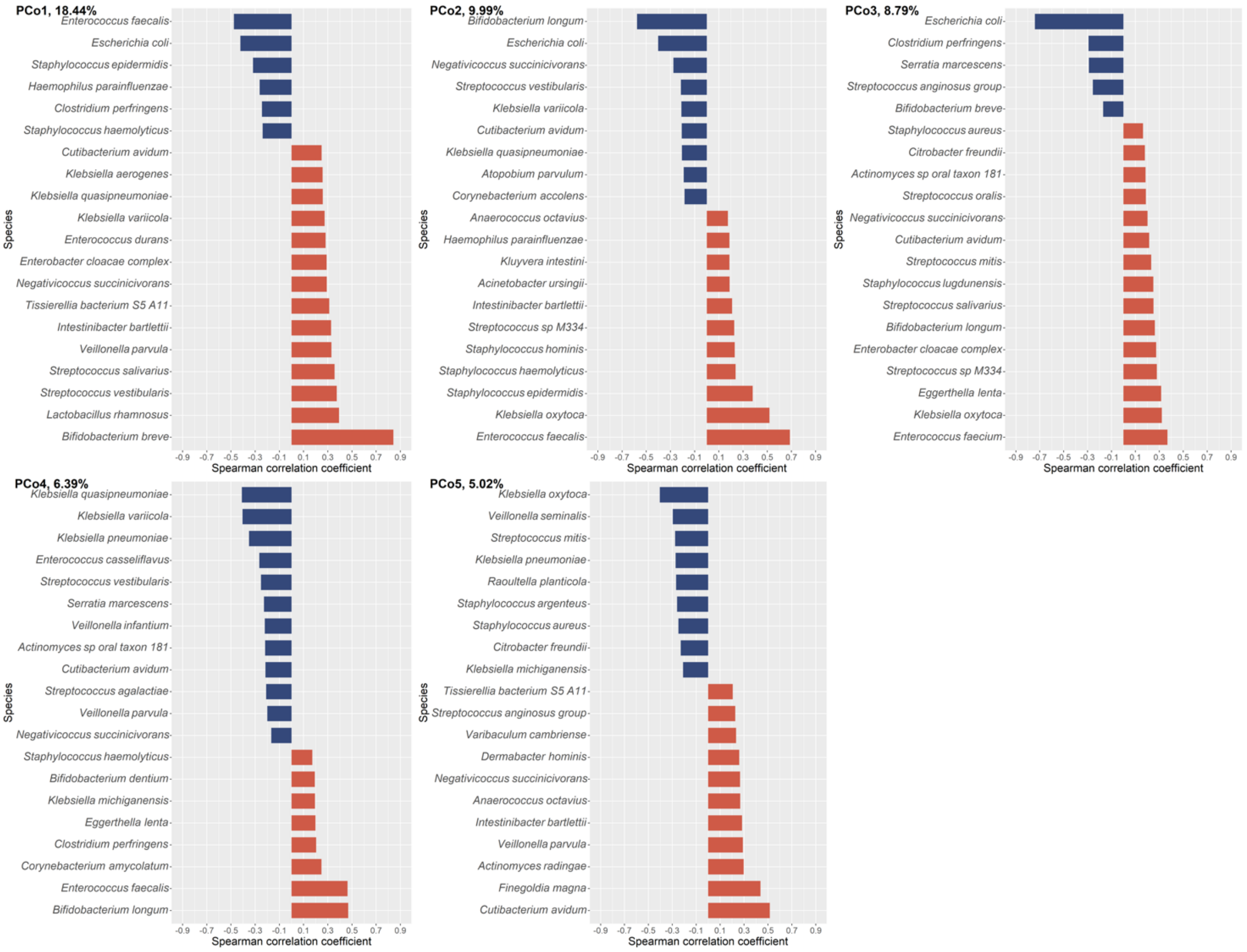


***Supplementary Figure 2. Dimensionality reduction of the microbiota community composition data. Bacterial species correlations with the first five orthogonal principal coordinates (PCo), derived from the Bray-Curtis dissimilarity matrix calculated on the relative abundances of bacterial species, showing the top 20 strongest correlations for each PCo. The % refers to the variance explained by each of the PCos. Red indicates positive and blue negative correlations between the PCo-s and species relative abundances.***

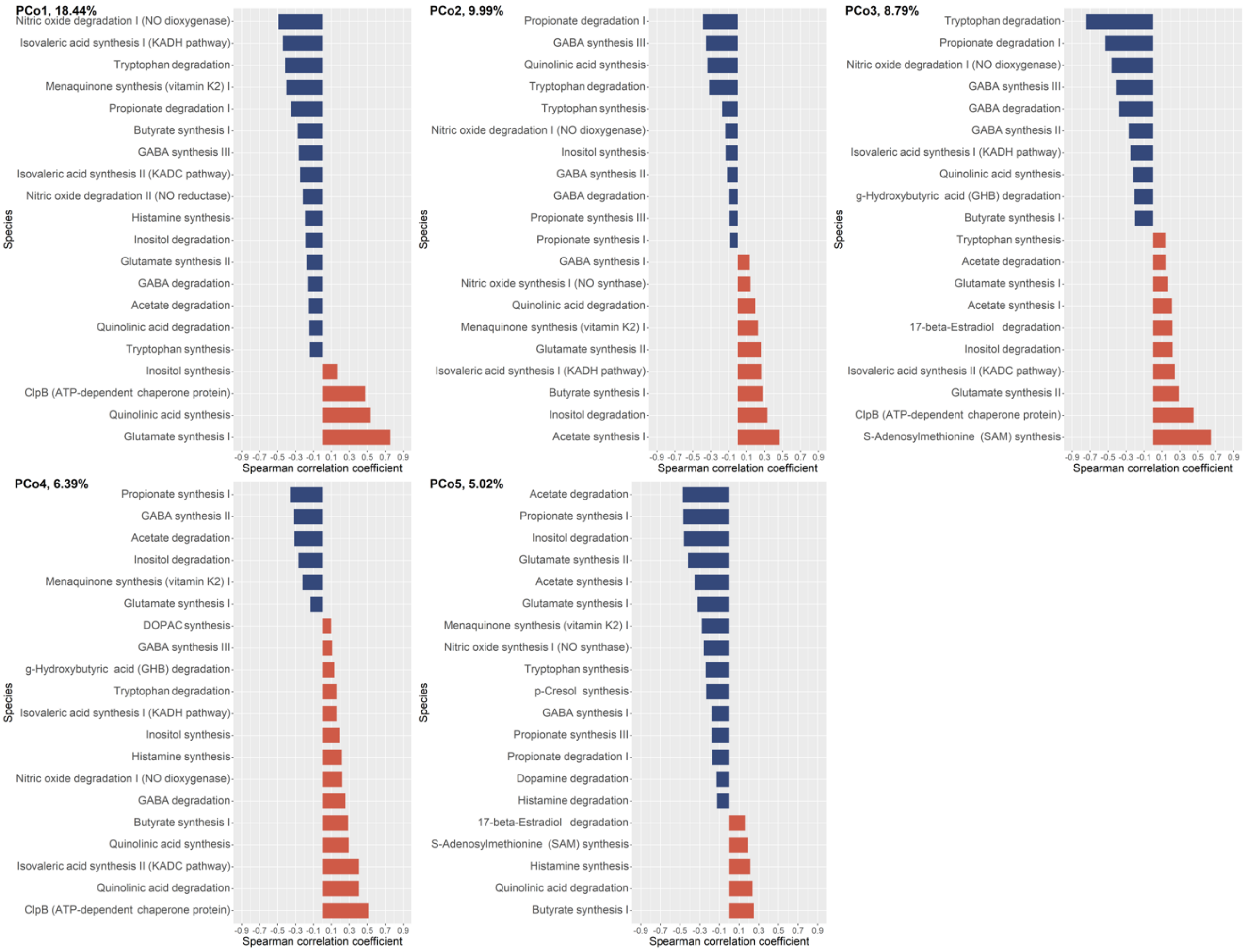


***Supplementary Figure 3. Gut-brain modules (GBM) abundance correlations with the five community composition principal coordinates (PCo), derived from the Bray-Curtis dissimilarity matrix calculated on the relative abundances of bacterial species, showing the top 20 strongest correlations for each PCo. The % refers to the variance explained by each of the PCos. Red indicates positive and blue negative correlations between the PCo-s and GBM relative abundances.***

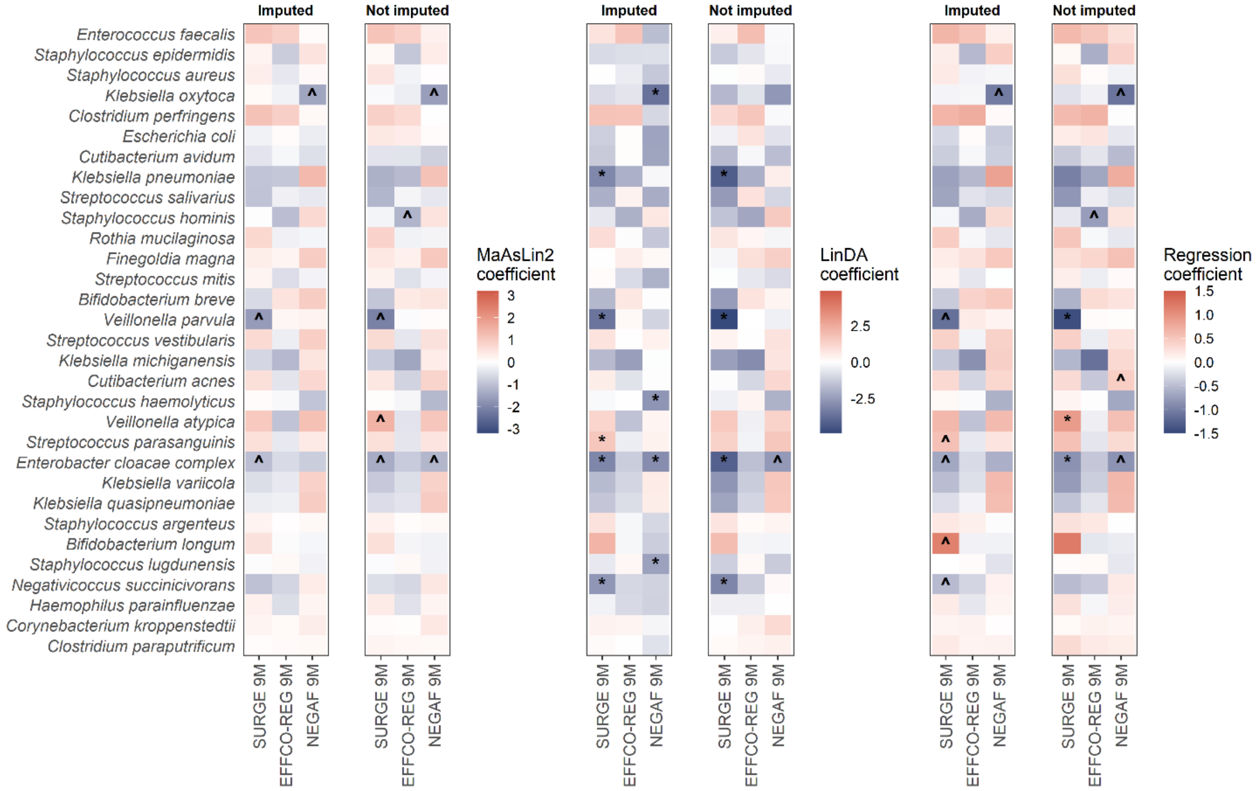


***Supplementary Figure 4. Gut microbiome species relative abundances in association with 9-month temperament data. The heatmaps show the results from the three differential abundance testing methods. Symbols denote significance level: *statistically significant (q < 0.05 after adjustment for multiple comparisons), ^nominally significant (p < 0.05 prior to adjustment). "Imputed" panels show results discussed in the main paper where the data was imputed for 10 infants; "Not imputed" panels show the results of the sensitivity analysis where those 10 infants with imputed data were excluded. There were minimal changes between the imputed vs non imputed results.***

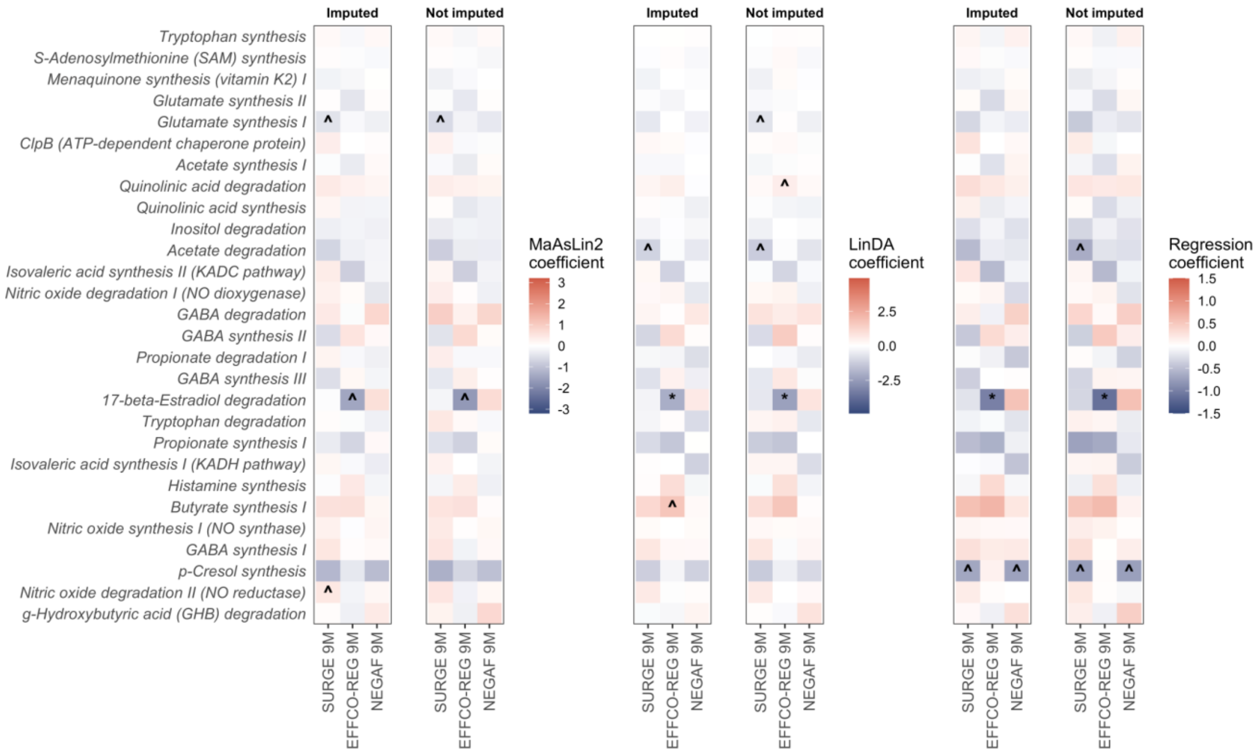


***Supplementary Figure 5. Gut-brain module relative abundances in association with 9-month temperament data. The heatmaps show the results from the three differential abundance testing methods. Symbols denote significance level: *statistically significant (q < 0.05 after adjustment for multiple comparisons), ^nominally significant (p < 0.05 prior to adjustment). "Imputed" panels show results discussed in the main paper where the data was imputed for 10 infants; "Not imputed" panels show the results of the sensitivity analysis where those 10 infants with imputed data were excluded. There were minimal changes between the imputed vs non imputed results.***

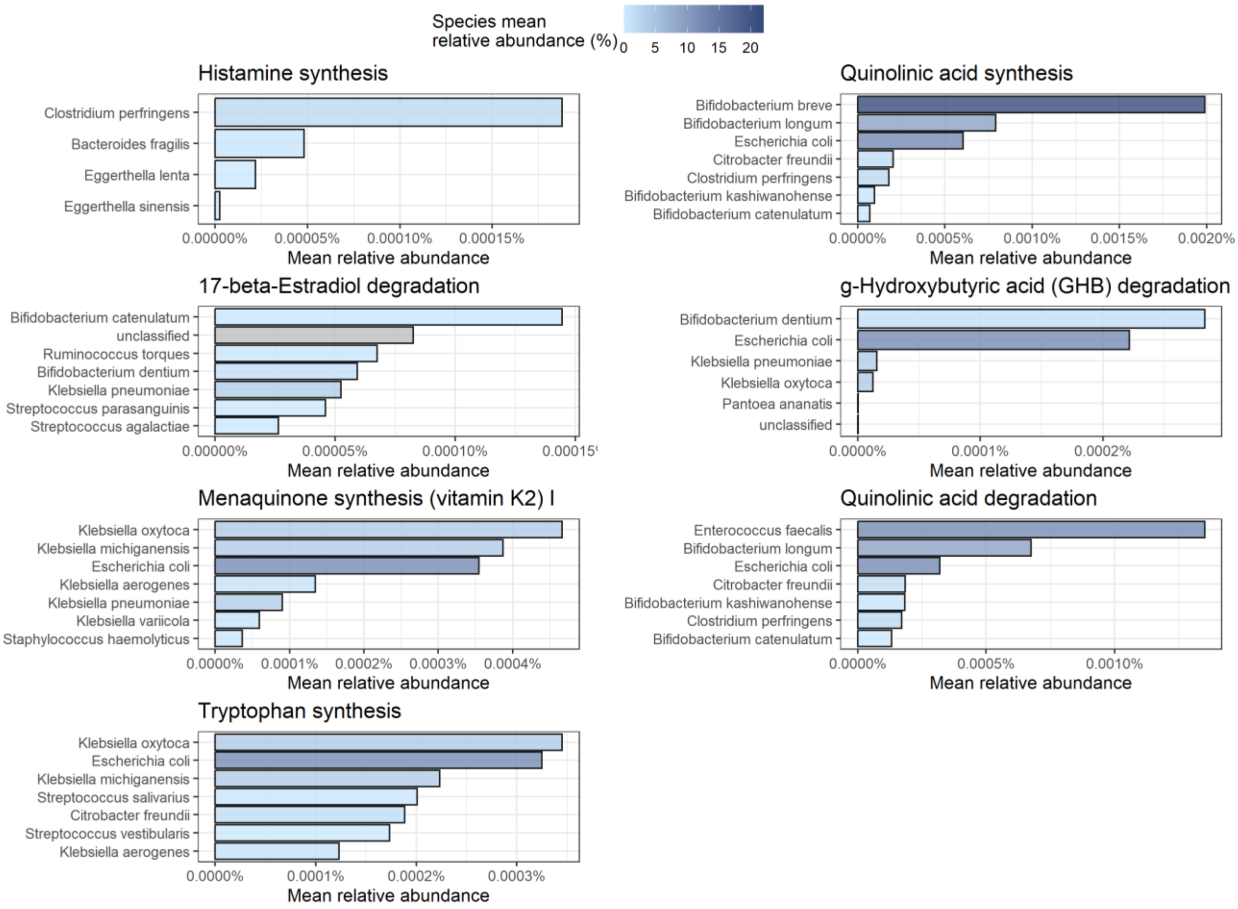


***Supplementary Figure 6. Species contribution to gut-brain modules with strong and moderate level evidence for associations with neurodevelopmental outcomes. Bar charts show the mean relative abundance of the gut-brain modules, stratified by species. Depth of the blue colour represents the mean relative abundance of the species in the dataset, with darker colours corresponding to higher overall relative abundance.***

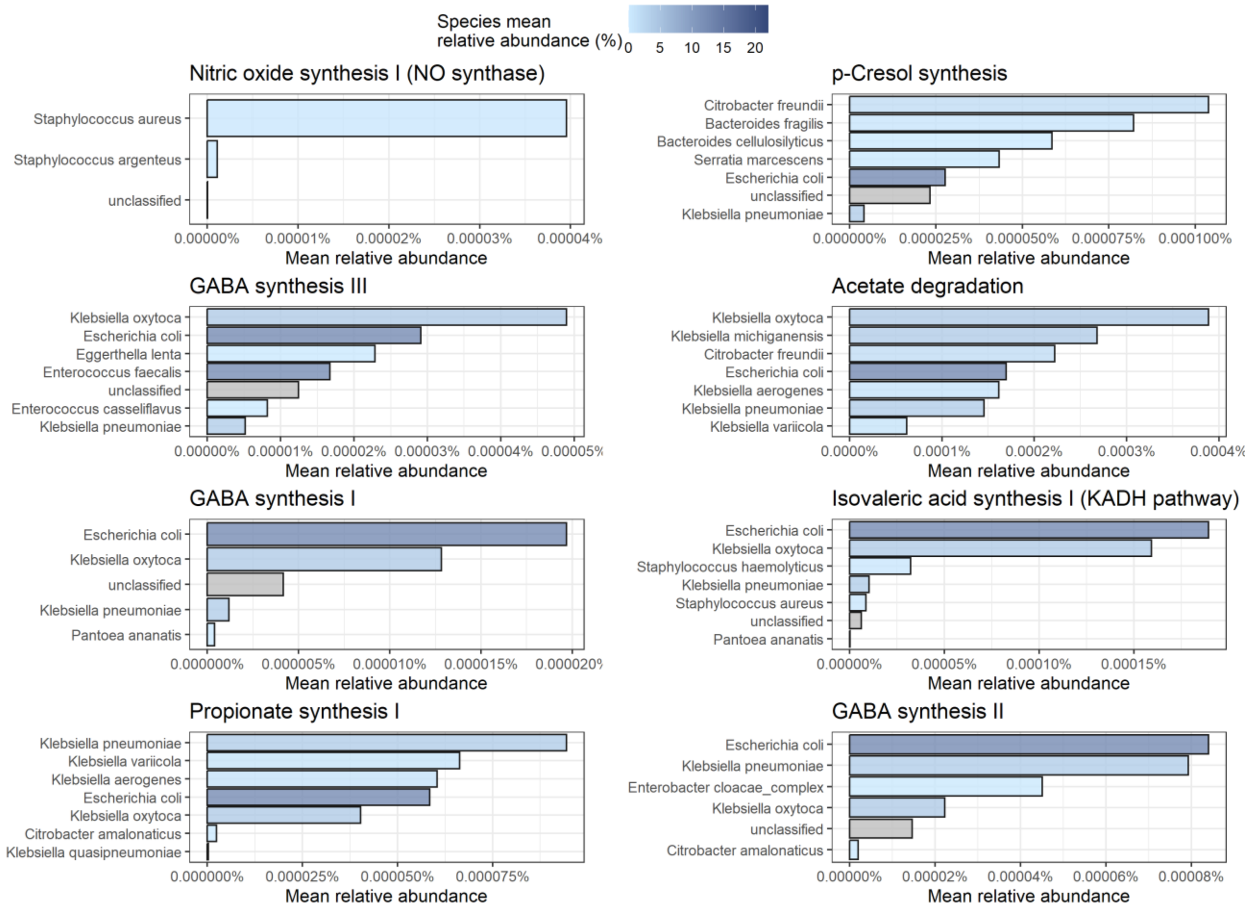


***Supplementary Figure 7. Species contribution to gut-brain modules with weak level evidence for associations with neurodevelopmental outcomes. Bar charts show the mean relative abundance of the gut-brain modules, stratified by species. Depth of the blue colour represents the mean relative abundance of the species in the dataset, with darker colours corresponding to higher overall relative abundance.***

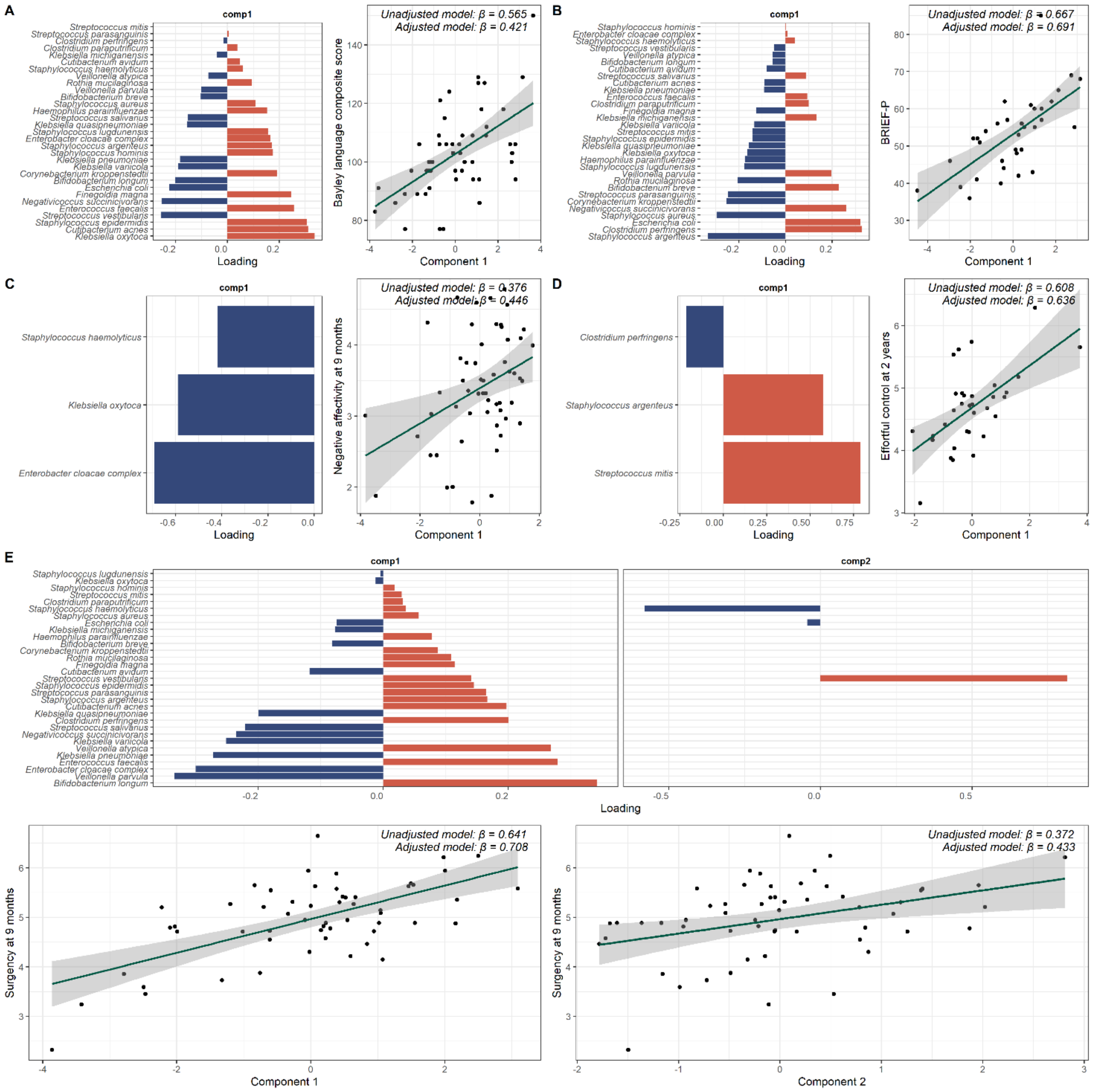


***Supplementary Figure 8. Sparse partial least squares analysis for species-level data, showing the loadings of species to the component(s) and correlation of the component(s) with the outcome measures for (A) Bayley language composite score, (B) BRIEF-P, (C) IBQ negative affectivity, (D) ECBQ effortful control and (E) IBQ surgency. Unadjusted β notes the standardised coefficient from the linear regression model associating the sPLS components with the outcome measures, adjusted β notes standardised coefficient from a linear regression model that additionally included GA at birth, PMA at sample, antibiotic exposure < 72h of life, birthweight z-score, high vs low proportion of exclusive breast milk days during NICU stay, infant sex, Scottish Index of Multiple Deprivation quintile, maternal BMI at pregnancy booking, and maternal age as covariates. IBQ surgency was the only outcome measure for which sPLS suggested a 2-component model.***

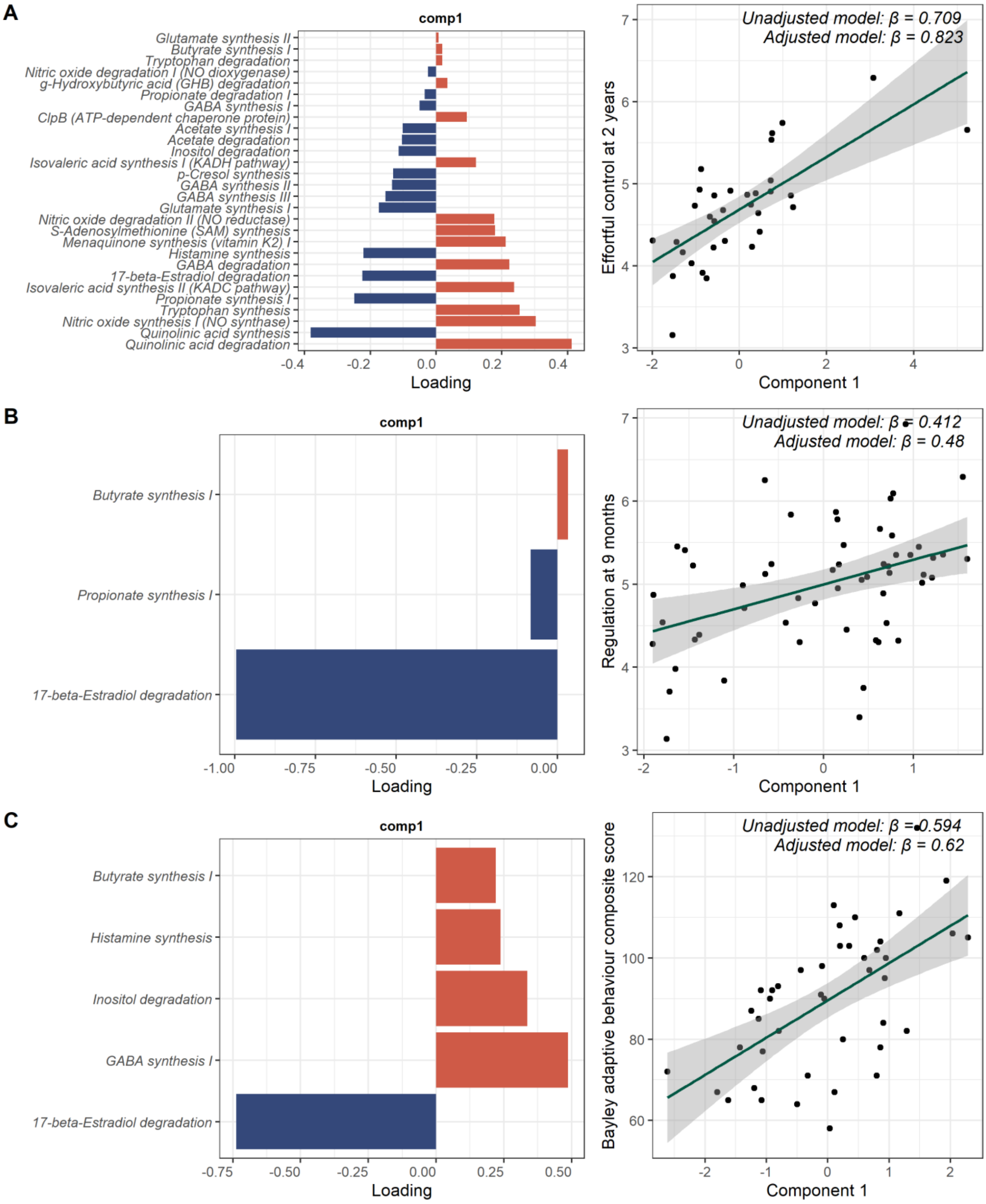


***Supplementary Figure 9. Sparse partial least squares analysis for gut-brain modules data, showing the loadings of gut-brain modules to the first component (left) and correlation of the first component with the outcome measures (right) for (A) ECBQ effortful control, (B) IBQ regulation, and (C) Bayley adaptive behaviour composite score. Unadjusted β notes the standardised coefficient from the linear regression model associating the PLS components with the outcome measures, adjusted β notes standardised coefficient from a linear regression model that additionally included GA at birth, PMA at sample, antibiotic exposure < 72h of life, birthweight z-score, high vs low proportion of exclusive breast milk days during NICU stay, infant sex, Scottish Index of Multiple Deprivation quintile, maternal BMI at pregnancy booking, and maternal age as covariates.***
